# Heterogeneity and heterotypic continuity of emotional and behavioural profiles across development

**DOI:** 10.1101/2019.12.26.19015917

**Authors:** João Picoito, Constança Santos, Carla Nunes

## Abstract

**Purpose:** To identify emotional and behavioural symptoms profiles from early childhood to adolescence, their stability across development and associated factors.

**Methods:** Our sample included 17,216 children assessed at ages 3, 5, 7, 11 and 14 years from the UK Millennium Cohort Study. We used latent profile and latent transition analysis to study their emotional and behavioural profiles from early childhood to adolescence. We included sociodemographic, family and parenting variables to study the effect on latent profile membership and transitions.

**Results:** The number and specific profiles of emotional and behavioural symptoms changed with the developmental stage. We found a higher number of profiles for ages 3, 5, and 14, suggesting greater heterogeneity in the presentation of emotional and behavioural symptoms in early childhood and adolescence compared to late childhood. There was greater heterotypic continuity between ages 3 and 5, particularly in transitions from higher to lower severity profiles. Children exposed to socioeconomic disadvantages were more likely to belong or transition to any moderate or high emotional and behavioural symptoms profiles. Maternal psychological distress and harsh parenting were associated with internalizing and externalizing profiles, respectively. Higher levels of internalizing and externalizing symptoms across development were associated with lower mental wellbeing and higher rates of self-harm and substance use in adolescence.

**Conclusion:** Emotional and behavioural symptoms develop early in life, with levels of heterogeneity and heterotypic stability that change throughout development. These results call for interventions to prevent and treat paediatric mental illness that consider the heterogeneity and stability of symptoms across development.

## Introduction

Emotional and behavioural (EB) symptoms often develop early in life and can have a lasting impact across development [1]. Children and adolescents with EB symptoms have higher odds of developing psychiatric disorders, poor academic achievement, and risk behaviours [2]. These enduring effects on the lives of young children have motivated research on the specific manifestations of early psychopathology. Identifying specific profiles of EB symptoms, and their transitions and associated factors is crucial for developmentally sensitive and effective interventions in childhood and adolescence mental health, within an ecological framework.

Traditionally, research on EB profiles in childhood and adolescence has focused on internalizing (such as depression and anxiety) and externalizing symptoms (such as hyperactivity, aggression and oppositional behaviour). Internalizing symptoms in childhood are associated with mood and anxiety disorders in adolescence and young adulthood [3], while externalizing symptoms are associated with conduct disorder and antisocial behaviours [4]. Although internalizing and externalizing symptoms are indeed distinct in terms of phenomenology, they tend to cluster together [5]. This warrants studying these manifestations with person-centred methods such as latent class or latent profile analysis (LPA) to capture different patterns of expression for internalizing and externalizing symptomatology, instead of studying these manifestations separately. Additionally, the different patterns of internalizing and externalizing symptoms may change from early childhood to adolescence as EB regulation skills develop, and new life challenges and difficulties arise.

EB symptoms can be transient or chronic, and have a variable course across development [3, 6]. Some children will develop similar problems (homotypic continuity), while others will develop different types of symptomatology (heterotypic continuity). The degree of homotypic or heterotypic stability and the factors that influence these transitions have been the topics of much research [7–9]. While homotypic stability has been reported as the norm [7], other studies have reported similar degrees of homotypic and heterotypic continuity [9]. It remains elusive whether some developmental periods are more prone to heterotypic or homotypic transitions, and which contextual and individual factors are associated with specific transitions.

In the current study, we wanted to know whether different developmental stages were associated with different degrees of heterogeneity in the expression of EB symptoms, and different degrees of homotypic and heterotypic continuity. Additionally, we wanted to explore which contextual and ecological factors had a greater influence on specific EB symptoms profiles and developmental stages. Thus, our main objectives were (1) to identify profiles of EB symptoms from early childhood to adolescence, and to study their transitions across development; (2) to assess the effect of socioeconomic, family and parenting factors on membership and transition to specific profiles; and (3) to explore the association between membership to specific EB symptoms profiles across development and mental wellbeing, self-harm, and substance use in adolescence. To achieve these objectives, we used person-centred statistical methods, namely LPA and latent transition analysis (LTA), on a large general population sample.

## Methods

### Participants

Data for this study were drawn from the Millennium Cohort Study (MCS; http://www.cls.ioe.ac.uk/mcs), an ongoing longitudinal study in the UK that has followed a sample of children since their infancy [10]. A stratified random sampling approach was used to overrepresent areas of high child poverty, and areas with high proportions of ethnic minorities. From the two initial sweeps (at age 9 months and 3 years, respectively), 19,243 families in total were enrolled in the MCS. Subsequent sweeps were conducted at ages 3, 5, 7, 11 and 14 years that included 15,590, 15,246, 13,857, 13,287, and 11,726 families, respectively. Additional details on study design, sampling, attrition, and ethics can be found elsewhere [11]. For the analysis, we included cohort members (singletons and first-born twins or triplets) with data on EB symptoms in at least one of sweeps 2 to 6, comprising 17,216 of 19,243 participants. Children excluded from the analysis (n=2,027) were more likely to be male, from ethnic minorities, living in a monoparental family, and from lower-income households (supplementary materials Table S1). Table 1 reports the sample characteristics.

**Table 1.** Weighted descriptive statistics (n= 17216)

| **Table 1 Weighted descriptive statistics (n= 17216)** |  |  |
| --- | --- | --- |
|  | n (%) /<br>mean (standard error) | Missing<br>n (%) |
| **Gender (male)** | 8804 (51.1) | - |
| **Ethnicity (white)** | 14138 (87.8) | 100 (0.6) |
| **Mother's professional qualification**<br>(high – National Vocation Qualification level<br>4 or higher vs. other) | 4963 (34.6) | 702 (4.1) |
| **Family income poverty**<br>(equivalized income <60% of the national<br>median) |  |  |
| Age 3 | 5053 (26.1) | 1880 (10.9) |
| Age 5 | 5115 (27.1) | 2118 (12.3) |
| Age 7 | 4155 (23.2) | 3396 (19.7) |
| Age 11 | 3298 (17.6) | 3949 (22.9) |
| Age 14 | 3457 (21.3) | 5511 (32) |
| **Monoparental family** |  |  |
| Age 3 | 2691 (15.3) | 1817 (10.6) |
| Age 5 | 3014 (17.9) | 2028 (11.8) |
| Age 7 | 2912 (19.2) | 3378 (19.6) |
| Age 11 | 3129 (22.2) | 3949 (22.9) |
| Age 14 | 2881 (23.5) | 5511 (32.0) |
| **SDQ Emotional problems** |  |  |
| Age 3 | 1.31 (0.17) | 2473 (14.4) |
| Age 5 | 1.35 (0.02) | 2490 (14.5) |
| Age 7 | 1.49 (0.02) | 3773 (21.9) |
| Age 11 | 1.82 (0.03) | 4421 (25.7) |
| Age 14 | 1.97 (0.03) | 5889 (34.2) |
| **SDQ Conduct problems** |  |  |
| Age 3 | 2.75 (0.03) | 2446 (14.2) |
| Age 5 | 1.46 (0.02) | 2471 (14.4) |
| Age 7 | 1.34 (0.02) | 3744 (21.7) |
| Age 11 | 1.34 (0.02) | 4418 (25.7) |
| Age 14 | 1.35 (0.02) | 5887 (34.2) |
| **SDQ Hyperactivity/inattention score** |  |  |
| Age 3 | 3.86 (0.03) | 2584 (15) |
| Age 5 | 3.23 (0.03) | 2557 (14.9) |
| Age 7 | 3.30 (0.04) | 3794 (22.0) |
| Age 11 | 3.05 (0.04) | 4445 (25.8) |
| Age 14 | 2.90 (0.04) | 5894 (34.2) |
| **SDQ Peer problems score** |  |  |
| Age 3 | 1.48 (0.02) | 2570 (14.9) |
| Age 5 | 1.09 (0.02) | 2500 (14.5) |
| Age 7 | 1.17 (0.02) | 3764 (21.9) |
| Age 11 | 1.32 (0.02) | 4414 (25.6) |
| Age 14 | 1.68 (0.03) | 5884 (34.2) |
| **SDQ Prosocial score** |  |  |
| Age 3 | 7.34 (0.02) | 2549 (14.8) |
| Age 5 | 8.39 (0.02) | 2472 (14.4) |
| Age 7 | 8.60 (0.02) | 3740 (21.7) |
| Age 11 | 8.78 (0.02) | 4411 (25.6) |
| Age 14 | 8.32 (0.02) | 5886 (34.2) |
| **Maternal distress (Kessler-6)** |  |  |
| Age 3 | 3.14 (0.04) | 3627 (21.1) |
| Age 5 | 3.07 (0.04) | 2885 (16.8) |
| Age 7 | 3.03 (0.05) | 4053 (23.5) |
| Age 11 | 3.87 (0.06) | 5511 (32) |
| Age 14 | 4.16 (0.07) | 6849 (39.8) |
| **Harsh parenting** (sum of 3 items from<br>Straus's Conflict Tactics Scale [smacks,<br>shouts and telling off]) |  |  |
| Age 3 | 9.39 (0.03) | 4220 (24.5) |
| Age 5 | 8.37 (0.02) | 3073 (17.8) |
| Age 7 | 8.11 (0.02) | 4203 (24.4) |

Results are presented as n (%) for categorical variables, and as mean (standard error) for continuous variables. n – unweighted frequency. % - weighted percentage. SDQ – Strengths and Difficulties Questionnaire

### Measures

The cohort members’ *EB profiles* were measured at ages 3, 5, 7, 11 and 14 with the parent-reported Strengths and Difficulties Questionnaire (SDQ), a brief behavioural screening questionnaire for children aged 3–16 years [12]. It is composed of 25 items, scored 0–2 (‘not true’, ‘somewhat true’ or ‘certainly true’), and divided into five subscales: emotional symptoms, conduct problems, hyperactivity and inattention, peer relationship problems and prosocial behaviour. Scores for each subscale range from 0–10, with higher scores indicating more problems or difficulties. Reliability and cut-offs used for interpretation of the profiles can be found in the supplementary materials (S1A).

The sociodemographic and family variables included gender, ethnicity (white vs. other), living in a monoparental family, family income poverty, maternal professional qualification, maternal psychological distress, and harsh parenting. More details on these variables can be found in the supplementary materials (S1B). We also assessed functional outcomes measured at age 14, namely mental wellbeing, self-harm and lifetime substance use, including alcohol, binge drinking, tobacco and cannabis.

### Data analysis

To assess EB profiles from early childhood to adolescence, we conducted the analysis in several steps.

First, we performed a cross-sectional LPA on the SDQ subscale scores and determined separately for each time-point the number of profiles. LPA is a type of finite mixture modelling that identifies discrete groups of individuals within a population, in which the latent variable is categorical, and the indicators are treated as continuous [13]. For each age we included all cases that had data for at least one SDQ subscale. The sample sizes included for each age were as follows: age 3 n=14,830; age 5 n=14,768; age 7 n=13,486; age 11 n=12,817; age 14 n=11,336. Missing values for the SDQ subscales were dealt with full-information maximum likelihood (FIML) procedures incorporated in LPA, assuming to be missing at random. To select the optimal number of latent profiles we used different criteria, namely the Bayesian information criterion (BIC), with lower values indicating better fit; the Lo-Mendell-Rubin test, which tests if an additional profile improves model fit; and entropy for each model, measured on a 0 to 1 scale, with values approaching 1 indicating less classification error; as well as interpretability, clinical meaningfulness and parsimony. More details on these fit indices can be found elsewhere [14].

Second, to study the transitions between profiles from early childhood to adolescence we performed LTA, a longitudinal extension of LPA that allowed us to calculate the transition probability from a profile at time t to another profile at time t+1 [13]. It is common in LTA to test for full measurement invariance, i.e. whether profiles are equal across time, because it allows for a straightforward interpretation of latent profiles across time [14]. However, as we were interested in capturing developmental changes in EB profiles from childhood to adolescence, measurement non-invariance was conceptually more appropriate than full measurement invariance. Additionally, full measurement invariance could mask significant developmental differences from childhood to adolescence [15]. Therefore, we tested for partial measurement invariance, which is a middle ground between full measurement invariance and measurement non-invariance, implicating equality constraints for some of the measurement parameters, but not all [15]. These models were compared with a model with measurement non-invariance, using the BIC, with lower values indicating better fit.

Third, we included covariates in the LTA model to assess the effect of socioeconomic, family and parenting factors on membership to specific EB symptoms profiles. Each covariate was adjusted for gender and ethnicity.

Fourth, we explored the influence of socioeconomic, family and parenting factors on how the children transitioned between EB symptoms profiles. This was achieved allowing for the covariates (one per model) to have an interaction on the transition probabilities, as described in [16].

For all LTA models, we used the manual three-step specification [16]. This method has two major advantages: preventing a profile at time t from influencing the formation of another profile at time t+1, and vice-versa; and avoiding measurement parameter shifts with the inclusion of covariates in the LTA model.

The manual three-step specification involved the estimation of the unconditional mixture model (LPA), the assignment of each participant to latent profiles using modal class assignment, and then estimating the models with measurement parameters fixed at values that accounted for the measurement error in the class assignment. A final LTA model was estimated using the three-step variables obtained earlier for each LPA model combined, including all time points. The LTA models included all participants with at least one latent profile assignment, comprising a total of 17,216. Missing latent profile assignments were dealt with FIML procedures incorporated in LTA.

Fifth, we explored the association between membership to specific EB symptoms profiles across development and mental wellbeing, self-harm, and substance use in adolescence. For this, we performed a series of models, regressing each adolescence outcome on age-specific EB symptoms profiles separately, adjusting for sociodemographic, family and parenting factors.

Missingness on the covariates cannot be treated with FIML approaches. Therefore, we multiply imputed by chained equations 25 datasets, using a model comprising all covariates and the SDQ subscale scores, and other variables related to the missing covariates. All analyses accounted for the stratified clustered sample design of the MCS. Data analysis was performed with Mplus version 8.3 [17], and the packages MplusAutomation [18] and dotwhisker [19] for R version 3.5.1. Mplus annotated syntax and excerpts of the outputs are available in the supplementary materials (S9).

## Results

### Latent profile analysis of EB symptoms

A series of LPA models were fit separately for each age, with models ranging from one to six classes. At ages 3, 5, and 14 a five-profile solution was considered the best fitting model, while for ages 7 and 11 a four-profile model provided a better fit. More details on the fit indices and rationale for the choice of LPA models can be found in the supplementary materials (S2).

Figure 1 shows the different latent profiles from early childhood to adolescence.

**Figure 1.**
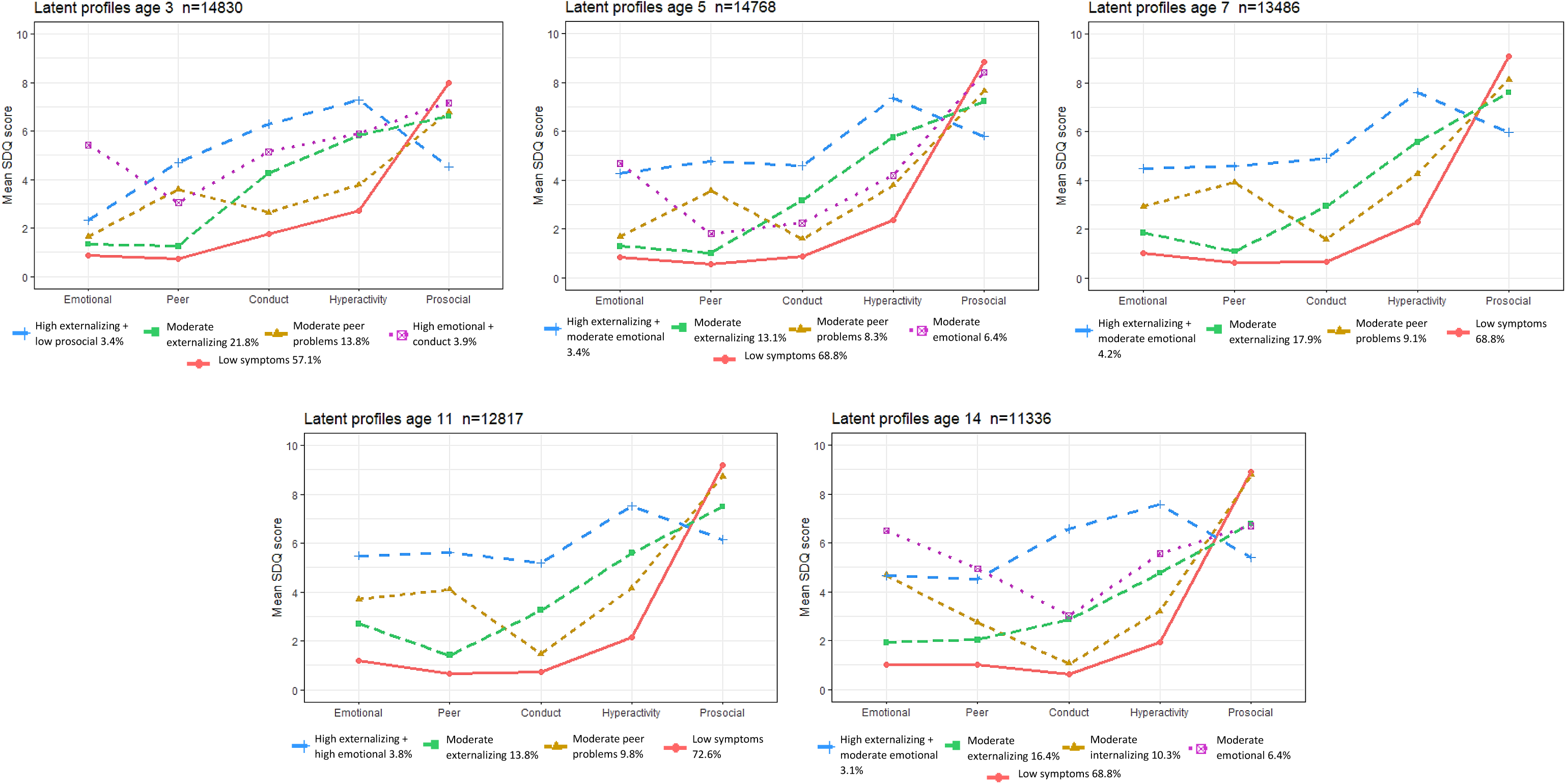
Latent profiles of emotional and behavioural symptoms.

#### Similar profiles found across all ages

Across all time points, we found a profile characterized by low scores on the symptoms scales and high prosocial behaviour (*low symptoms)* comprising the majority of the sample (57.1% at age 3, and peaking at 72.6% at age 11). The second most common profile was *moderate externalizing*, characterized by borderline scores on conduct problems and hyperactivity/inattention, with a stable shape from childhood to adolescence. From ages 3 through to 7, we found a profile of slightly raised peer relationship problems with average scores in the remaining subscales comprising 13.8, 8.3 and 9.1%, at ages 3, 5 and 7, respectively.

#### Profiles found at specific ages

At ages 11 and 14, a similar profile to *moderate peer problems* arose but with borderline scores on emotional problems and peer relationship problems named *moderate internalizing*. At age 14, additional to the *moderate internalizing* profile, we found a profile with higher scores for emotional and peer relationship problems *(high internalizing)*, with a prevalence of 5.1%. At age 3, we found a *high emotional and conduct* profile (3.9%) that was not found at age 5, when a *moderate emotional* profile appeared instead (6.4%). A profile of high externalizing symptoms (conduct problems and hyperactivity/inattention) emerged from childhood to adolescence, albeit with some distinct nuances. At age 3, this latter profile was associated with low prosocial behaviours *(high externalizing and low prosocial*, 3.8%), and at ages 5 and 7, it was instead associated with borderline emotional problems (*high externalizing and moderate emotional*, 3.4%, 4.2% and 3.1%, respectively). At age 11, this high externalizing profile was associated with higher emotional problems *(high externalizing and high emotional*, 3.8%).

#### Sensitivity analyses

We studied four-profile solutions for ages 3, 5, and 14 (Figures S1-S3) and found clinically meaningful differences compared to the chosen five-profile solutions, supporting our choice of five profiles for those ages.

As the sample size differed across time points, we examined whether this was responsible for the changes in the number and specificity of EB symptoms profiles across development. The sensitivity analyses, available in the supplementary materials (S4), did not provide evidence that missingness was responsible for the differences in latent profiles of EB symptoms across development, thus supporting our findings.

### Latent transition analysis of EB symptoms from childhood to adolescence

As we found an equal number of profiles between ages 3 and 5, and ages 7 and 11, we estimated a model with measurement non-invariance, and three models with partial measurement invariance (a model with equal profiles across ages 3 and 5; a model with equal profiles across ages 7 and 11; and a model with equal profiles across ages 3 and 5, and 7 and 11). The model with measurement non-invariance provided a better fit than the models with partial measurement invariance, evidenced by the lower BIC (supplementary materials S5). Figure 2 shows the transitions between latent profiles across time.

**Figure 2.**
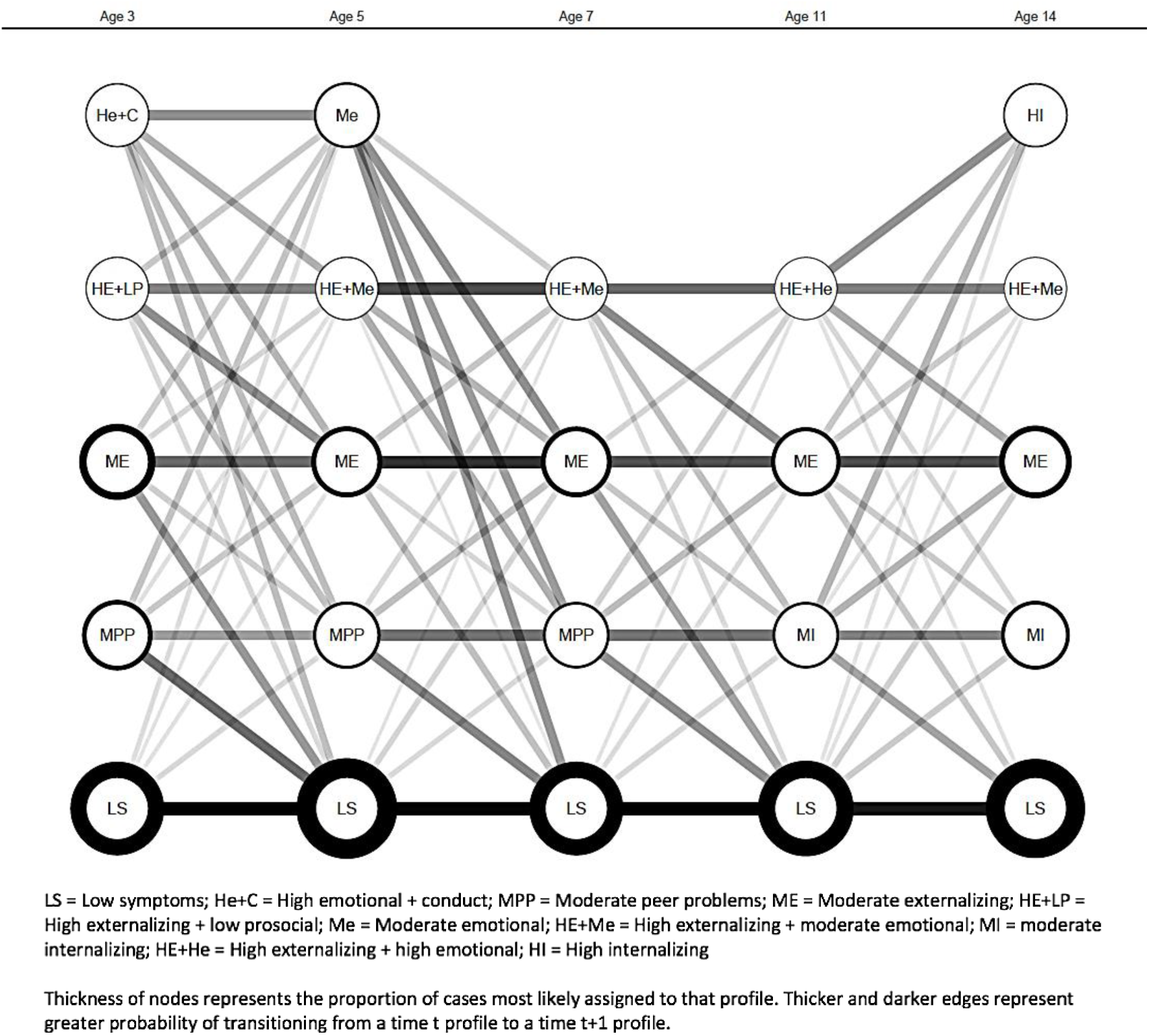
Latent transition probabilities and classification probabilities.

#### Profiles with high or increasing homotypic continuity

Children in the *low symptoms* profile had a high probability of remaining in the same profile at a later time-point, with transition probabilities of 93.2% (age 3 to 5) to 83.4% (age 11 to 14). Children in the *moderate externalizing* profile at age 3 were most likely to remain in the same profile at age 5 (48.4%) but also to transition to the *low symptoms* profile (31.1%). From age 5 onwards, the probability of staying in the *moderate externalizing* profile was higher, ranging from 58.0–74.3%, while the probability of transitioning to the *low symptoms* profile decreased (10.4–26.4%).

#### Profiles with high heterotypic continuity

The *moderate peer problems* profile, from ages 3 to 5, had a higher probability of transitioning to the *low symptoms* profile (50.5%) than staying in the same profile (27.5%). This pattern of a higher probability of transitioning to the *low symptoms* profile then changed in later transitions with a slightly higher probability of remaining in the same profile, and from ages 7–11 and 11–14 moving to the closely related *moderate internalizing* profile (44.4% and 35.7%, respectively). Children in the *high emotional and conduct problems* profile (specific to age 3) were likely to transition to the lower severity *moderate emotional* profile (38.4%), but also to others such as the *high externalizing and moderate emotional* (17.5%) or *low symptoms* profiles (14.5%). At age 3, those in the *high externalizing and low prosocial* profile had a slightly higher probability of transitioning to the related *high externalizing and moderate emotional* (38.3%) or *moderate externalizing* profiles (36.6%). From ages 5 to 7, children in the *high externalizing and moderate emotional* profile had a 65.3% probability of remaining in the same profile. This changed from age 7 to 11 with a lower probability of transitioning by age 11 to the closely related *high externalizing and high emotional* (47.5%) or the *moderate externalizing* profiles (34.7%). Children in the *high externalizing and high emotional* profile had a similar probability of transitioning to the related *high externalizing and moderate emotional* (38.4%) or the *high internalizing* profiles (33.2%).

### Socioeconomic and contextual differences

Figure 3 and Supplementary Tables S15-S19 present the results of the LTA with covariates. Boys were more likely to be in high and moderate externalizing profiles (except at ages 11 and 14) than girls, with higher odds for the age 3 *high externalizing and low prosocial* profile (odds ratio [OR]: 3.01, 95% confidence interval [CI] 2.43–3.74). White children were consistently less likely to belong to the *moderate peer problems* profile from ages 3 to 7 (age 3 OR: 0.49, CI: 0.41–0.6; age 5 OR: 0.53, CI: 0.42–0.67; age 7 OR: 0.54, CI: 0.42–0.69). Children from monoparental families and households with income below the poverty line were more likely to belong to any EB profile than to the *low symptoms* profile from ages 3 to 11, particularly at age 3 *high externalizing and low prosocial* (monoparental family OR: 3.68, CI: 2.89–4.68; poverty OR: 5.33, CI: 4.15–6.84) and *high emotional and conduct* (monoparental family OR: 2.81, CI: 2.23–3.54; poverty OR: 5.49, CI: 4.38–6.89). Similarly, adolescents living in households with income below the poverty line were more likely to belong to the *high externalizing and moderate emotional* profile (OR: 6.95, CI: 4.53–10.66). Children from mothers with a high professional qualification were less likely to belong to any moderate or high EB profile, especially at age 3 *high emotional and conduct* (OR: 0.24, CI: 0.18-0.34). Higher maternal distress was consistently associated with a higher risk of inclusion in any EB symptoms profile, compared to the *low symptoms* profile at any age. Harsh parenting was associated with higher odds of membership in any externalizing profile at any age, but particularly at age 7 *(high externalizing and moderate emotional* OR: 1.75, CI: 1.59–1.93; *moderate externalizing* OR: 1.62, CI: 1.52–1.73).

**Figure 3.**
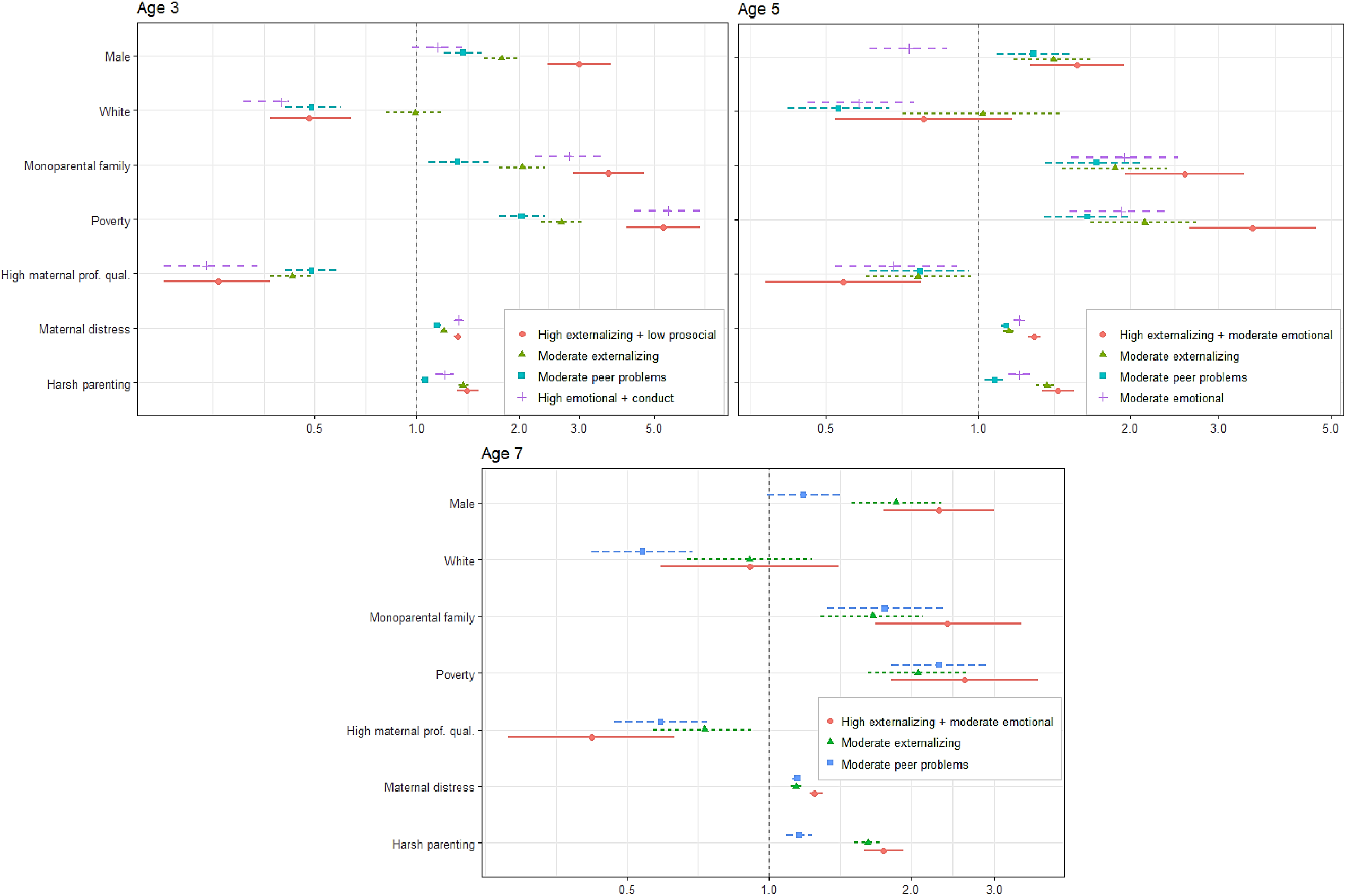
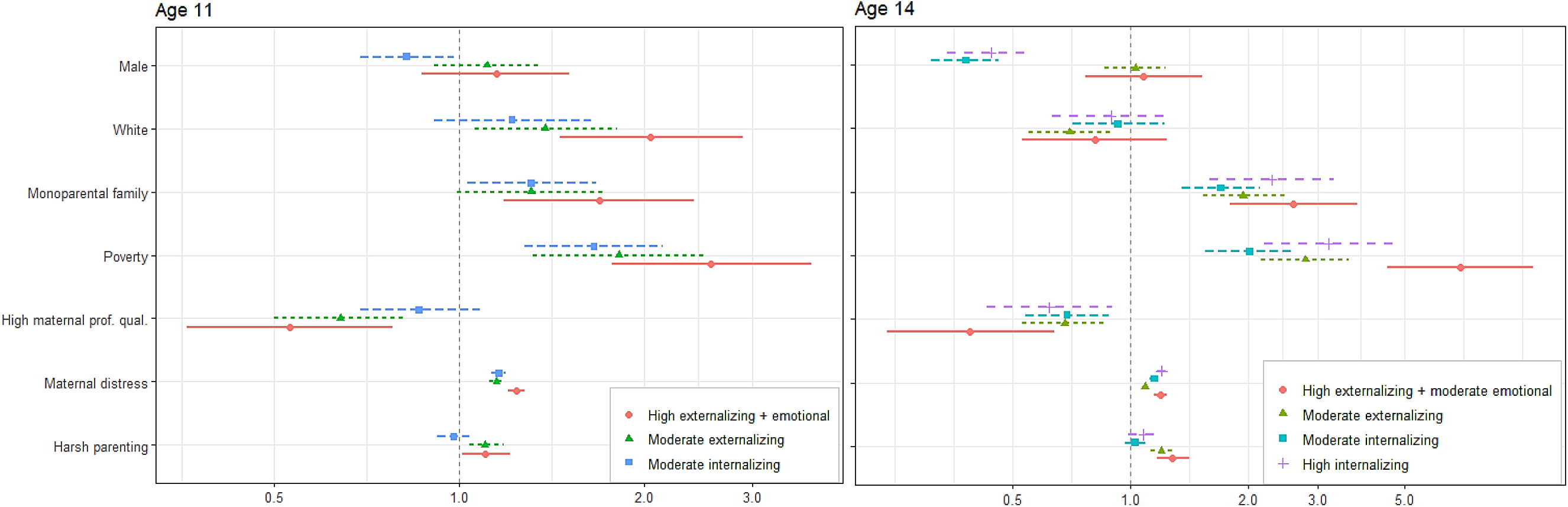
Results of the latent transition analysis with covariates. Monoparental family (time-varying), poverty (time-varying), high maternal professional qualification, maternal distress (time-varying) and harsh parenting (time-varying) odds ratios are adjusted for gender and ethnicity. Reference class = low symptoms.

### Influence of covariates on the transition probabilities

We additionally tested whether contextual covariates modified the probability of a child transitioning among profiles (Supplementary Tables S20–S33). Children from monoparental families and households with income poverty were more likely to transition to higher severity profiles, internalizing or externalizing, at any developmental stage (for example, age 3 *moderate externalizing* to age 5 *high externalizing and moderate emotional* profile: poverty OR: 1.79, CI: 1.06–3.01, monoparental family OR: 2.49, CI: 1.29-4.79). Harsh parenting was associated with maintenance of or transition to moderate externalizing profiles such as age 5 *moderate externalizing* to age 7 *high externalizing and moderate emotional* (OR: 1.23, CI: 1.04–1.47), or age 5 *moderate peer problems* to age 7 *high externalizing and moderate emotional* (OR: 1.41, CI: 1.15–1.73). Maternal distress was consistently associated with higher odds of transitioning to moderate or high internalizing profiles, such as age 3 *low symptoms* to age 5 *moderate emotional* (OR: 1.11, CI: 1.03–1.21), age 7 *low symptoms* to age 11 *moderate internalizing* (OR: 1.19, CI: 1.07–1.17), age 11 *moderate internalizing* to age 14 *high internalizing* (OR: 1.13, CI: 1.07–1.20). High maternal professional qualification was associated with a lower risk of transitioning to moderate or high severity profiles, such as age 5 *low symptoms* to age 7 *moderate externalizing* (OR: 0.6 CI: 0.40–0.89), age 7 *low symptoms* to age 11 *moderate internalizing* (OR: 0.57, CI: 0.39–0.85), and age 11 *low symptoms* to age 14 *moderate externalizing* (OR: 0.60, CI: 0.44–0.83).

### Association between EB symptoms profiles and adolescent mental wellbeing, self-harm, and substance use (Table 2)

Compared to the *low symptoms* profile, children and adolescents from any EB profile (except for age 3 *moderate peer problems, and high emotional and conduct)* were more likely to report lower mental wellbeing in adolescence, especially for the high externalizing profiles. Ages 5 and 7 *high externalizing and moderate emotional*, and *moderate externalizing* profiles were associated with higher odds of reporting self-harm in adolescence. For ages 11 and 14, all EB profiles were associated with self-harm, especially age 14 *high externalizing and moderate emotional* (OR: 4.11, CI: 3.01–5.62) and *high internalizing* (OR: 3.45, CI: 2.70–4.42). Children and adolescents in the age 5, 7, 11 and 14 *moderate externalizing* profiles had higher odds of reporting in adolescence positive alcohol, binge drinking, smoking and cannabis lifetime use. Age 3 *moderate externalizing* was also associated with smoking (OR: 1.30, CI: 1.12-1.53) and cannabis use (OR: 1. 34, CI: 1.03-1.75) in adolescence. Contrastingly, ages 3 and 5 *moderate peer problems* were associated with a lower risk of a positive lifetime alcohol use in adolescence.

**Table 2.** Association between EB symptoms profiles and adolescence mental wellbeing, self-harm, and substance use.

| Latent profiles<br>(ref. Low<br>symptoms) | Mental<br>wellbeing<br>Coeff. (95%<br>CI) | Self-harm<br>OR (95% CI) | Lifetime<br>alcohol<br>use<br>OR (95% CI) | Lifetime<br>binge<br>drinking<br>OR (95% CI) | Lifetime<br>smoking<br>OR (95% CI) | Lifetime<br>cannabis<br>use<br>OR (95% CI) |
| --- | --- | --- | --- | --- | --- | --- |
| Age 3 |  |  |  |  |  |  |
| High externalizing<br>and low prosocial | **-1.52**<br>**(-2.37, -0.68)** | 1.08<br>(0.75, 1.54) | **0.64**<br>**(0.50, 0.84)** | 0.98<br>(0.61, 1.57) | 1.25<br>(0.89, 1.75) | 0.94<br>(0.53, 1.68) |
| Moderate<br>externalizing | **-0.72**<br>**(-1.13, -0.31)** | 1.11 (0.94,<br>1.31) | 1.02<br>(0.92, 1.14) | 1.12<br>(0.94, 1.34) | **1.30**<br>**(1.12, 1.53)** | **1.34**<br>**(1.03, 1.75)** |
| Moderate peer<br>problems | -0.29<br>(-0.70, 0.13) | 1.05<br>(0.87, 1.26) | **0.77**<br>**(0.67, 0.87)** | 0.84<br>(0.68, 1.04) | 0.91<br>(0.76, 1.08) | 0.75<br>(0.54, 1.04) |
| High emotional and<br>conduct | -0.09<br>(-0.76, 0.57) | 0.96<br>(0.69, 1.31) | **0.68**<br>**(0.53, 0.87)** | 1.01<br>(0.67, 1.53) | 1.19<br>(0.90, 1.59) | 1.13<br>(0.71, 1.80) |
| Age 5 |  |  |  |  |  |  |
| High externalizing<br>and moderate<br>emotional | **-1.60**<br>**(-2.42, -0.79)** | **1.39**<br>**(1.01, 1.92)** | **0.65**<br>**(0.50, 0.85)** | 1.06<br>(0.69, 1.62) | **1.43**<br>**(1.04, 1.98)** | 0.98<br>(0.59, 1.63) |
| Moderate<br>externalizing | **-0.80**<br>**(-1.21, -0.39)** | **1.27**<br>**(1.06, 1.52)** | 1.14<br>(0.99, 1.31) | **1.34**<br>**(1.10, 1.64)** | **1.40**<br>**(1.18, 1.66)** | **1.43**<br>**(1.07, 1.91)** |
| Moderate peer<br>problems | **-1.04**<br>**(-1.59, -0.50)** | 1.01<br>(0.79, 1.29) | **0.77**<br>**(0.65, 0.91)** | 0.85<br>(0.64, 1.13) | 1.03<br>(0.84, 1.27) | 1.02<br>(0.73, 1.43) |
| Moderate emotional | **-0.80**<br>**(-1.40, -0.21)** | 1.09<br>(0.85, 1.40) | **0.73**<br>**(0.59, 0.90)** | 0.96<br>(0.70, 1.33) | 0.99<br>(0.79, 1.25) | 0.90<br>(0.58, 1.39) |
| Age 7 |  |  |  |  |  |  |
| High externalizing<br>and moderate<br>emotional | **-2.29**<br>**(-3.13, -1.47)** | **1.79**<br>**(1.31, 2.45)** | 0.97<br>(0.75, 1.24) | 1.23<br>(0.58, 1.78) | **1.87**<br>**(1.38, 2.55)** | **1.65**<br>**(1.04, 2.62)** |
| Moderate<br>externalizing | **-0.99**<br>**(-1.40, -0.59)** | **1.37**<br>**(1.16, 1.61)** | **1.15**<br>**(1.03, 1.29)** | **1.36**<br>**(1.12, 1.66)** | **1.77**<br>**(1.50, 2.10)** | **1.85**<br>**(1.42, 2.40)** |
| Moderate peer<br>problems | **-0.99**<br>**(-1.51, -0.47)** | 1.22<br>(0.97, 1.53) | 0.85<br>(0.72, 1.00) | 0.96<br>(0.77, 1.22) | 1.09<br>(0.88, 1.35) | 1.08<br>(0.73, 1.60) |
| Age 11 |  |  |  |  |  |  |
| High externalizing<br>and high emotional | **-3.12**<br>**(-4.02, -2.23)** | **2.39**<br>**(1.70, 3.37)** | 0.84<br>(0.64, 1.10) | 0.99<br>(0.64, 1.52) | **1.84**<br>**(1.31, 2.57)** | **1.86**<br>**(1.11, 3.10)** |
| Moderate<br>externalizing | **-1.96**<br>**(-2.42, -1.50)** | **1.49**<br>**(1.23, 1.79)** | **1.26**<br>**(1.09, 1.47)** | **1.6**<br>**(1.30-1.98)** | **2.10**<br>**(1.77, 2.49)** | **2.21**<br>**(1.68, 2.91)** |
| Moderate<br>internalizing | **-1.89**<br>**(-2.44, -1.34)** | **1.62**<br>**(1.32, 1.98)** | **0.77**<br>**(0.66, 0.90)** | 0.91<br>(0.69, 1.21) | 0.88<br>(0.68, 1.13) | 0.98<br>(0.64, 1.50) |
| Age 14 |  |  |  |  |  |  |
| High externalizing<br>and moderate<br>emotional | **-5.37**<br>**(-6.26, -4.49)** | **4.11**<br>**(3.01, 5.62)** | **1.86**<br>**(1.42, 2.44)** | **3.11**<br>**(2.17, 4.47)** | **5.57**<br>**(4.15, 7.47)** | **7.06**<br>**(4.78, 10.43)** |
| Moderate<br>externalizing | **-2.11**<br>**(-2.51, -1.70)** | **1.69**<br>**(1.42, 2.01)** | **1.42**<br>**(1.25, 1.63)** | **1.88**<br>**(1.56, 2.26)** | **2.44**<br>**(2.09, 2.84)** | **2.65**<br>**(2.07, 3.40)** |
| Moderate<br>internalizing | **-2.54**<br>**(-3.03, -2.04)** | **2.19**<br>**(1.79, 2.69)** | 0.88<br>(0.75, 1.03) | 0.99<br>(0.77, 1.28) | 1.00<br>(0.79, 1.28) | 0.99<br>(0.61, 1.63) |
| High internalizing | **-4.52**<br>**(-5.25, -3.79)** | **3.45**<br>**(2.70, 4.42)** | 0.91<br>(0.73, 1.14) | **1.43**<br>**(1.04, 1.97)** | **2.06**<br>**(1.60, 2.66)** | **2.51**<br>**(1.65, 3.81)** |

Results adjusted for gender, ethnicity, poverty (at 9 months of age), monoparental family (at 9 months of age), high maternal professional qualification (at 9 months of age), harsh parenting (at age 3), and maternal distress (at age 3),

## Discussion

We used LPA and LTA to capture developmental changes in EB profiles from childhood to adolescence. We found that the number and specific profiles of EB symptoms that could best explain heterogeneity in the data changed with the developmental stage. Early childhood was characterized by profiles with higher internalizing and externalizing symptoms, compared with later stages. We found the emergence at age 5 of a profile with mixed externalizing and internalizing symptomatology that persisted until adolescence. We found increased levels of internalizing symptoms at ages 11 and 14. The higher number of profiles for ages 3, 5 and 14 suggests increased heterogeneity in the presentation of EB difficulties in early childhood and adolescence compared with late childhood and preadolescence (ages 7 and 11).

Our study builds on the findings of a recent study [5] that also examined EB symptoms from age 3 to 11, using data from the MCS. The authors found five different groups of EB symptoms for ages 3 to 11 based on the conduct problems and emotional problems subscales of the SDQ [5]. The five subgroups were similar to our results, namely low symptoms (57% of the sample), moderate behavioural (21%), moderate emotional (12.5%), high emotional and moderate behavioural (5.5%), and high behavioural and moderate emotional classes (4%) [5]. A recent LTA study on problematic behaviour described four profiles of problem behaviour at ages 1.5, 3 and 6, including an internalizing and externalizing profile, that displayed higher scores for aggressive behaviour and dysregulation at age 6 [8]. The finding of a mixed internalizing and externalizing profile present across development points to a dynamic interplay between these two constructs [20].

### Homotypic and heterotypic continuity changes throughout development

Compared to later developmental stages, more heterotypic continuity was observed between ages 3 and 5, particularly transitions from high to moderate profiles and to the *low symptoms* profile. The decline in externalizing symptoms from ages 3 to 5 has been reported in the literature and is explained by the typical maturation and development of cognition and affect regulation processes [21, 22]. In contrast, homotypic transition was the rule from ages 5 to 11, with a consistently high probability of remaining in the same profile. A community study of childhood psychiatric disorders in Norway found higher homotypic continuity between 8 and 10 years than between 4 and 6 years [7]. Another previous study on childhood psychiatric disorders between ages 3 and 6, reported a similar magnitude of homotypic and heterotypic stability in this age range [9]. Past early childhood, EB problems seemingly tended to crystallize and stabilize, with fewer children transitioning to the *low symptoms* profile. Indeed, early childhood is considered a developmentally sensitive period that is particularly permeable to protective and risk factors [23, 24]. The transition to adolescence was also associated with a higher degree of heterotypic continuity, particularly to internalizing profiles and to lower prevalence in the *low symptoms* profile. Accordingly, adolescence has been associated with increased rates of internalizing disorders [25] and with homotypic and heterotypic pathways to these disorders [26].

### Socioeconomic disadvantage is associated with higher severity and chronicity of EB symptoms

Our analysis included a set of socioeconomic and contextual covariates to study the effects of these factors on membership to specific EB profiles, and to assess the effect on transition probabilities. Living in a monoparental family and household poverty were associated with a higher probability of membership and transition to any moderate or high EB profile. These odds were particularly high in early childhood. A systematic review on socioeconomic inequalities and mental health among children and adolescents reported that socioeconomic hardship was more strongly associated with mental health problems among younger children than among adolescents [27]. Single parenthood has been linked with a range of functional and mental health problems in both children and adolescents [28, 29]. This association may stem from the higher probability of economic hardship, mental illness in parents, and adverse life events in single-parent families [30]. Maternal distress was associated with membership in any moderate or high EB problems profile compared to the *low symptoms* profile and consistently associated with higher odds of transition to an internalizing profile. Maternal psychological distress and mental illness have been considered strong predictors of children’s mental ill-health and adjustment, and thus an important focus for treatment and prevention of paediatric mental illness [31]. Children exposed to harsh parenting practices were more likely to belong or transition to externalizing profiles (including mixed internalizing and externalizing). The association between externalizing symptoms and harsh parenting has been consistently replicated in the literature, particularly within a transactional theoretical framework [32]. A recent study using data from the MCS found that harsh parenting measured at age 3 was associated with fewer conduct problems but higher emotional problems at age 11 [33]. The methodological differences from our study could explain the contrasting results; namely, we used a person-centric approach and included harsh parenting as a time-varying covariate (measured at ages 3, 5 and 7). However, the frequent co-occurrence of internalizing and externalizing symptoms (also evidenced in our results for the *high externalizing and moderate emotional* profile, for example) could also explain these differences. Children of mothers with high professional qualifications had higher odds of belonging to the low symptoms profile, particularly from ages 3 to 7. Low maternal education has been associated with both mental illness in children and adolescents and its persistence and severity [34], with higher effects for younger children [35].

### Early EB symptoms are associated with worse outcomes in adolescence

We studied the association between EB profiles across development and mental wellbeing, self-harm and substance use in adolescence. These associations helped validate the profiles’ clinical meaningfulness. Children and adolescents in the *high externalizing and moderate emotional*, and *moderate externalizing* profiles had the highest likelihood of reporting positive lifetime use of alcohol, binge drinking, smoking and cannabis, as well as self-harm and lower mental wellbeing. These associations were found even for age 3 and 5 externalizing profiles. The link between internalizing and externalizing symptoms, mental wellbeing, and risk behaviours in adolescence may stem from the behavioural and emotional self-control and regulation that develops early in life [36].

### Strengths and limitations

To our best knowledge, this is the first study to assess EB profiles from early childhood to adolescence within an LTA framework that includes five time points. We have included both time-invariant and time-varying covariates and studied their effect on the transition probabilities. Although computationally heavy, these models allowed us to capture developmental changes from childhood to adolescence. We used the five subscales of the SDQ and allowed for more nuanced EB profiles that could not have been achieved using just a subset of the subscales. Additional strengths are the large sample provided by the MCS and its national representativeness.

One important limitation to consider is the somewhat subjective choice of number of profiles in LPA. We used different fit criteria and applied them coherently in our analysis to overcome this limitation. Additionally, the subjective interpretation of the latent profiles must be considered when reading our results. Another limitation is the use of only parent-reporting for the SDQ subscales, which can produce some bias in the results due to discrepancies in the reporting of internalizing and externalizing symptoms between different sources (parent, teacher and child) [37]. The EB profiles do not represent clinical disorders. However, the SDQ has shown good predictive power for clinical diagnoses [12]. The higher attrition rate for socioeconomically disadvantaged families and those from ethnic minorities in each sweep of the MCS might have biased our results. However, we tried to minimize this, incorporating weights in the analyses that account for attrition and non-response.

### Conclusion

EB symptoms tend to develop early in life, with levels of heterogeneity, and heterotypic stability that change with the developmental stage. There is more heterogeneity in the presentation of EB difficulties in early childhood and adolescence, than in late childhood. Past early childhood, EB problems tend to crystallize and stabilize, with fewer children transitioning to the low symptoms profile. Children from socioeconomically disadvantaged families were more likely to belong to moderate or high internalizing or externalizing profiles, while high maternal professional qualification was protective, particularly for younger children. Higher levels of internalizing and externalizing symptoms, even in early childhood, were associated with lower mental wellbeing and higher rates of self-harm and substance use later on. Thus, within an ecological framework, earlier intervention for EB symptoms is crucial for better mental health in children and adolescents. These results highlight the need to implement developmentally sensitive interventions for the prevention and treatment of mental disorders in children and adolescents that consider the heterogeneity and stability of symptoms across development.

## Data Availability

The authors are grateful to the Centre for Longitudinal Studies (CLS), UCL Institute of Education, for the use of these data and to the UK Data Service for making them available. However, neither CLS nor the UK Data Service bear any responsibility for the analysis or interpretation of these data.

https://beta.ukdataservice.ac.uk/datacatalogue/series/series?id=2000031

## Funding

The authors have no funding to report.

## Conflict of interest

On behalf of all authors, the corresponding author states that there is no conflict of interest.

## SUPPLEMENTARY MATERIALS

### S1 - SUPPLEMENTARY MATERIALS - METHODS

#### S1A. Strengths and Difficulties Questionnaire reliability and cut-offs used

We assessed the reliability of the SDQ subscales by computing ordinal coefficient alpha, which is considered a suitable alternative to Cronbach’s alpha for ordinal scales, particularly for scales with few response items [1, 2]. The reliability was good for all SDQ subscales with ordinal alpha ranging across sweeps at 0.76–0.82 for emotional symptoms, 0.65–0.8 for peer relationship problems, 0.78–0.86 for hyperactivity and inattention, 0.77–0.85 for conduct problems, and 0.77–0.86 for prosocial behaviour. Each subscale was used in the analysis as continuous variables. To interpret the severity, we used proposed cut-off points by Goodman, based on British norms [3] (5 for emotional problems, 4 for conduct and peer problems, 7 for hyperactivity/inattention, and 4 for prosocial behaviour) [4]. At age 3, the cut-off point was lower for emotional problems (4) and higher for conduct problems (5) and prosocial behaviour (7) [5].

#### S1B. Sociodemographic and family variables details

The choice of covariates was based on the existing literature [6–8]. We included in the analysis time-invariant and time-varying covariates.

##### Time-invariant covariates

- Gender
- Ethnicity (white vs. other)
- Maternal professional qualification at 9 months of age for cohort member (National Vocational Qualification level 4 or higher vs. other) [9].

##### Time-varying covariates

- Maternal psychological distress was measured with the self-reported Kessler K6 [10] (Cronbach’s alpha 0.87–0.89), measured at ages 3, 5, 7, 11 and 14.
- Harsh parenting was assessed with the sum of three items from Straus’s Conflict Tactics Scale [11], (Cronbach’s alpha 0.66–0.67). These items measure how often a parent uses violent physical or verbal discipline practices such as smacks, shouts and “telling off,” with higher values indicating a higher frequency [12], measured at ages 3,5 and 7.
- Living in a monoparental family. Measured at ages 3, 5, 7, 11 and 14.
- Family income poverty (equivalized income <60% of the national median). Measured at ages 3, 5, 7, 11 and14.

#### Functional outcomes measured at age 14

Mental wellbeing was measured using a six-item questionnaire (Cronbach’s alpha = 0.86) in which adolescents reported their level of happiness with various aspects of their lives, namely family, school, appearance, and life overall.

For self-harm, adolescents were asked “Have you hurt yourself on purpose in any way in the past year?”

**Table S1.**
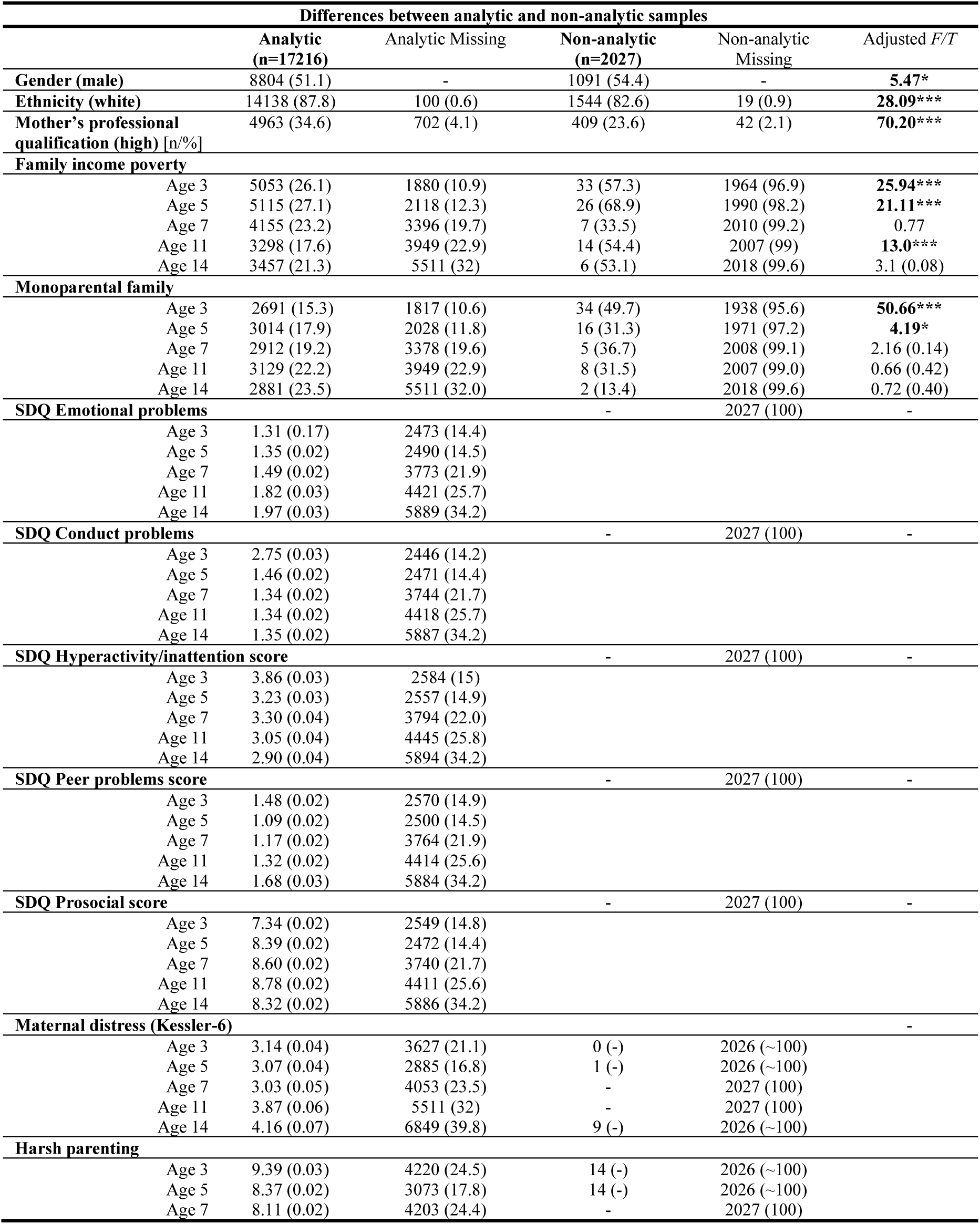
Differences between analytic and non-analytic samples

#### S2. Latent profile analysis of emotional and behavioural symptoms

A series of LPA models were fit separately for each age, with models ranging from one to six classes. At ages 3 and 5, a five-profile solution was considered the best fitting model, with the LMRT supporting the better fit of a five-profile compared to a four-profile solution, and with good entropy values (0.78 and 0.85 for ages 3 and 5, respectively). Although the six-profile solution presented better information criteria for ages 3 and 5, the LMRT p-value was not significant and entropy was lower at both ages compared to the five-profile solutions. At ages 7 and 11, solutions with six profiles also provided the lowest information criteria. However, the LMRT did not support a five- or six-profile solution, so we chose a four-profile solution for ages 7 and 11. Additionally, the entropy for the four-profile solutions was better than the nearest profiles for both ages 7 and 11. At age 14, the LMRT supported a five-profile solution with good entropy (0.85). Examining the resulting models for interpretability, clinical and developmental meaningfulness, and parsimony, we chose five-profile solutions for ages 3, 5, and 14, and four-profile solutions for ages 7 and 11. Tables S2-6 show the fit indices that supported model selection.

**Table S2.** Latent profile analysis fit indices (age 3)

| Number of classes | AIC | AICC | BIC | aBIC | Entropy | LRT p-value |
| --- | --- | --- | --- | --- | --- | --- |
| 2 | 288611.3 | 288611.3 | 288733.0 | 288682.1 | 0.762 | 0.0000 |
| 3 | 285691.2 | 285691.3 | 285858.5 | 285788.6 | 0.765 | 0.0000 |
| **4** | 284261.2 | 284261.3 | 284474.1 | 284385.1 | 0.784 | 0.0010 |
| **5** | **282741.7** | **282741.8** | **283000.2** | **282892.2** | **0.775** | **0.0000** |
| 6 | 281837.8 | 281838.1 | 282142.0 | 282014.9 | 0.774 | 0.5637 |

**Table S3.** Latent profile analysis fit indices (age 5)

| Number of classes | AIC | AICC | BIC | aBIC | Entropy | LRT p-value |
| --- | --- | --- | --- | --- | --- | --- |
| 2 | 272009.4 | 272009.4 | 272131.0 | 272080.2 | 0.861 | 0.0000 |
| 3 | 269029.8 | 269029.8 | 269197.0 | 269127.0 | 0.813 | 0.0395 |
| 4 | 266332.6 | 266332.8 | 266545.5 | 266456.5 | 0.844 | 0.0000 |
| **5** | **264766.5** | **264766.7** | **265024.9** | **264916.9** | **0.853** | **0.0000** |
| 6 | 263507.3 | 263507.5 | 263811.3 | 263684.2 | 0.847 | 0.6396 |

**Table S4.** Latent profile analysis fit indices (age 7)

| Number of classes | AIC | AICC | BIC | aBIC | Entropy | LRT p-value |
| --- | --- | --- | --- | --- | --- | --- |
| 2 | 253314.1 | 253314.1 | 253434.3 | 253383.4 | 0.874 | 0.0000 |
| 3 | 250065.3 | 250065.4 | 250230.6 | 250160.6 | 0.820 | 0.0047 |
| **4** | **247475.4** | **247475.5** | **247685.7** | **247596.7** | **0.858** | **0.0139** |
| 5 | 246029.2 | 246029.4 | 246284.5 | 246176.5 | 0.856 | 0.1085 |
| 6 | 244952.8 | 244953.1 | 245253.2 | 245126.1 | 0.862 | 0.7199 |

**Table S5.**
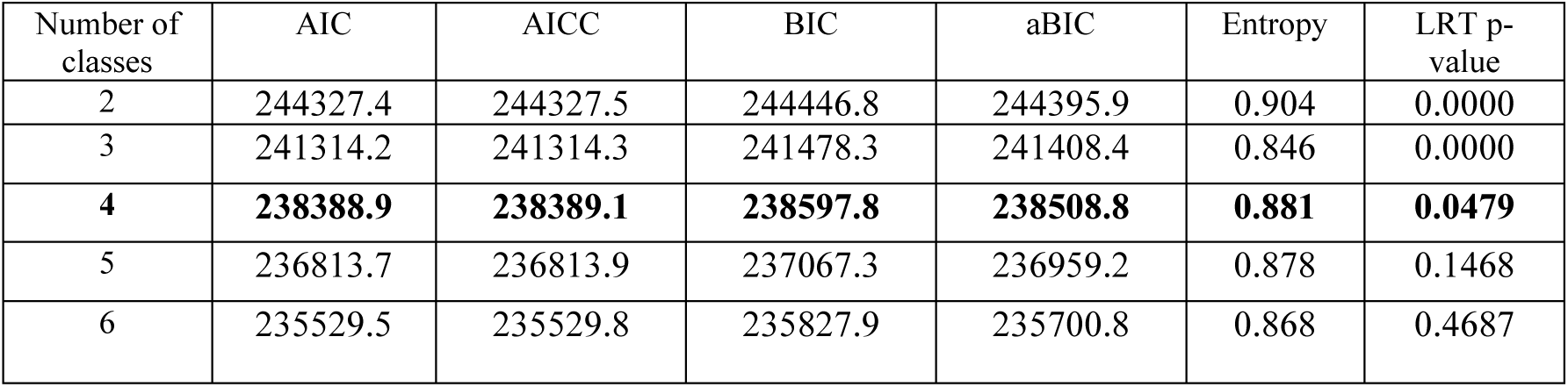
Latent profile analysis fit indices (age 11)

**Table S6.**
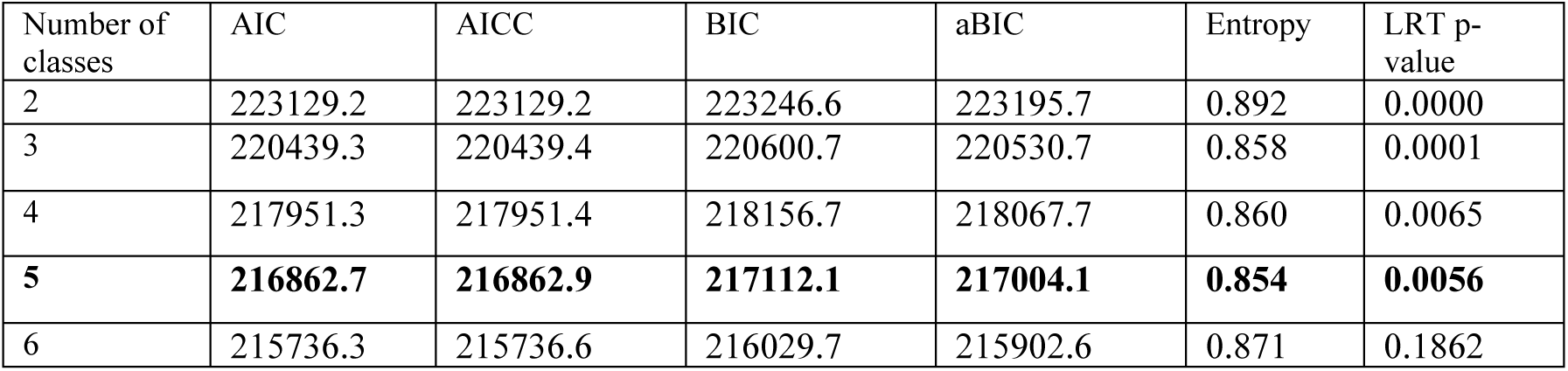
Latent profile analysis fit indices (age 14)

#### S3. Sensitivity analysis: Four-profile solutions for ages 3, 5, and 14

We also examined four-profile solutions for ages 3, 5, and 14 (figures S1-3) and found clinically meaningful differences compared to the chosen five-profile solutions. For example, the age 3 four-profile solution lacked the *high externalizing and low prosocial*, as well as the *moderate peer problems* profiles; the age 5 four-profile solution lacked the *moderate emotional* profile (comprising 6.4% of the children in the five-profile solution); age 14 four-profile solution lacked the *moderate internalizing* profile (10.3% of the adolescents in the five-profile solution).

**Figure S4.**
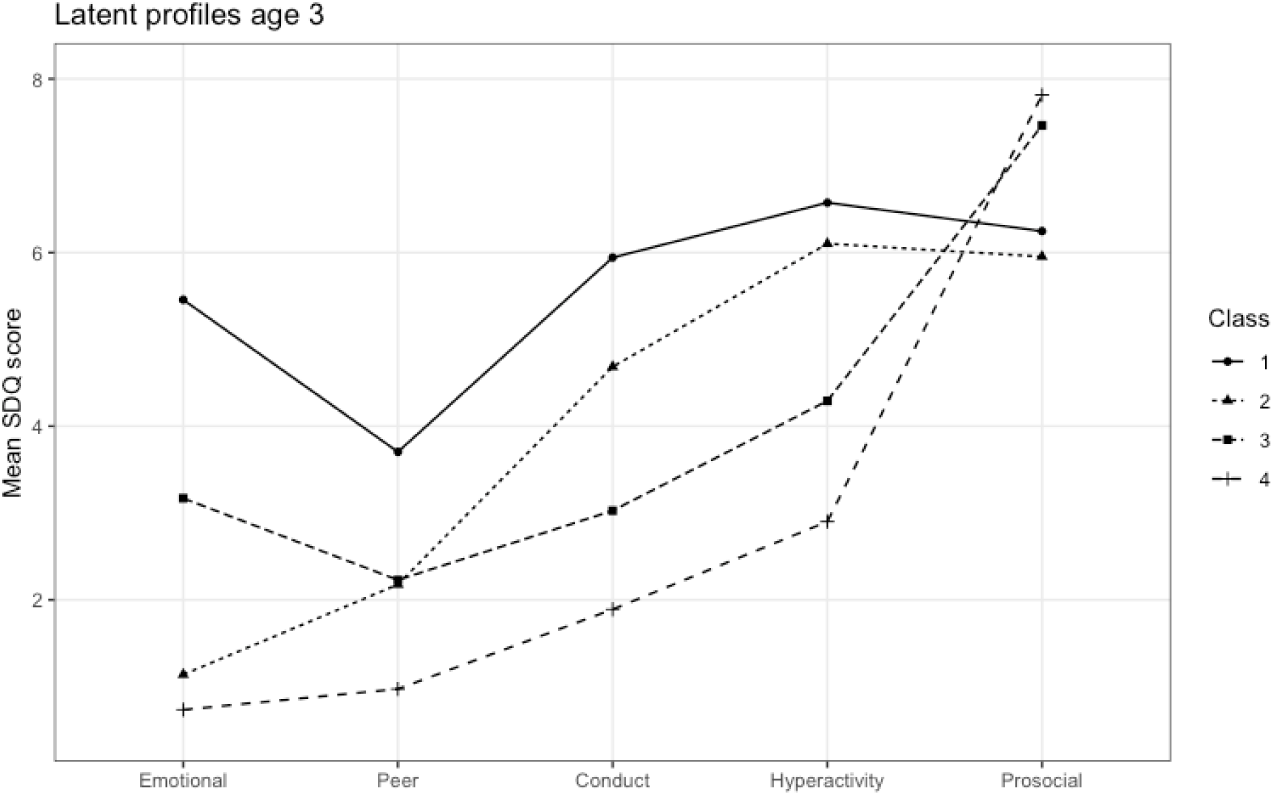
Age 3 four-profile model 1 - High emotional + conduct 2 - Subthreshold externalizing 3 - Subthreshold emotional 4 - Low symptoms

**Figure S5.**
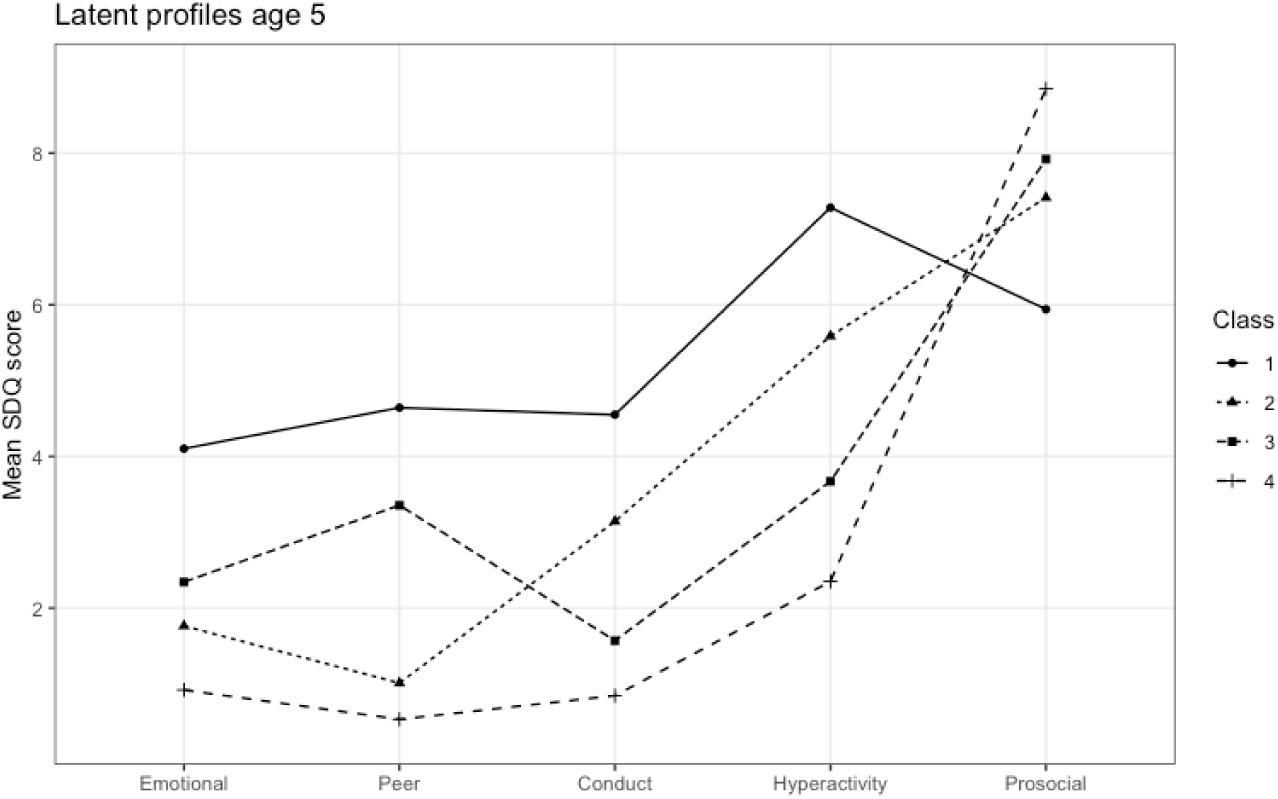
Age 5 four-profile model 1 - High externalizing + moderate emotional 2 - Moderate externalizing 3 - Moderate peer problems 4 - Low symptoms

**Figure S6.**
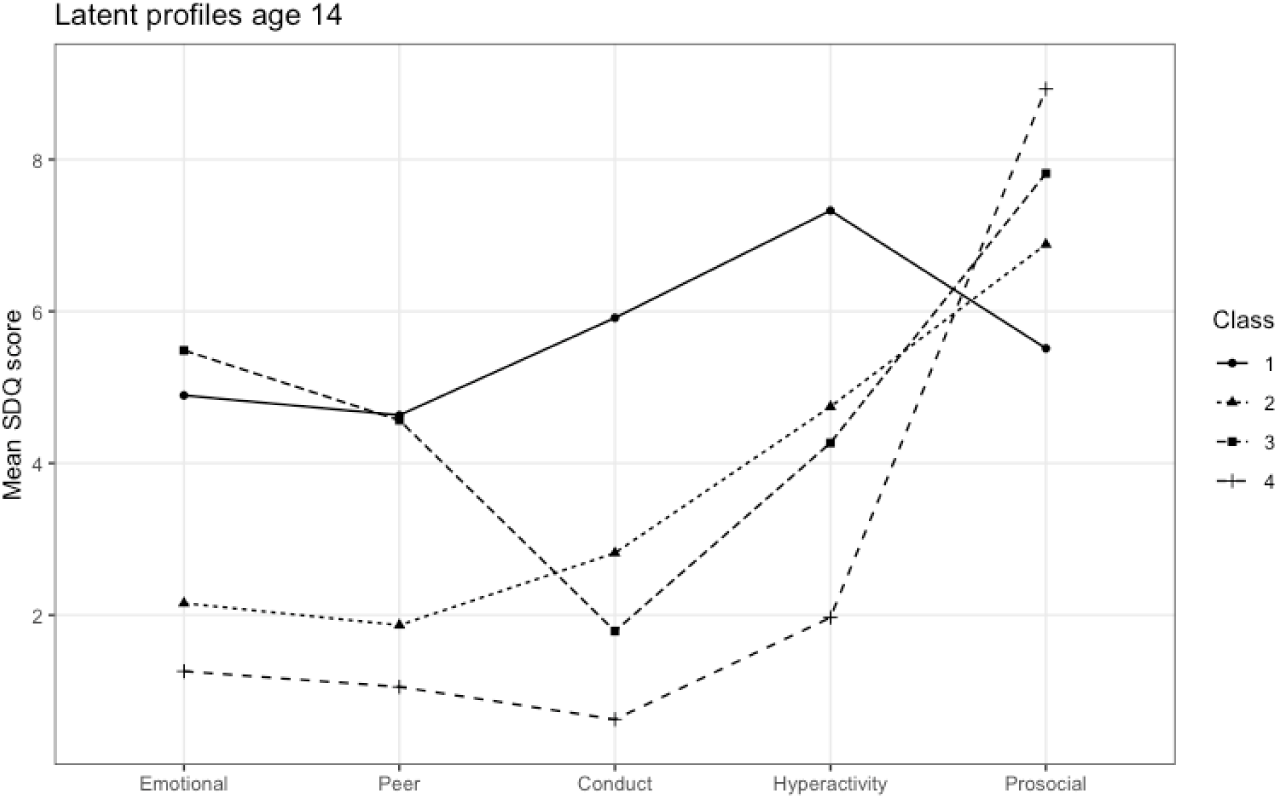
Age 14 four-profile model 1 - High externalizing + moderate emotional 2 - Moderate externalizing 3 - High externalizing 4 - Low symptoms

#### S4. Sensitivity analysis: influence of missing cases on latent profiles

##### Missing status at time t+1 and latent profile membership at time t

As the sample size differed across time points, we examined if this was responsible for the changes in number and specificity of profiles of EB symptoms across development. To accomplish this, we regressed being missing at time t+1 on latent profile membership at time t, controlling for gender, ethnicity, monoparental family, poverty and maternal professional qualification (Tables S7-10). We controlled for these variables because they are associated with attrition in the MCS [13]. At age 3, there was no association between latent profile membership and being missing at age 5. At age 5, children in the *high externalizing and moderate emotional* profile, compared to the *low symptoms* profile, were more likely to be missing at age 7. For ages 7 and 11, children in the *moderate externalizing* profile were more likely to be missing in the following sweeps. As the profiles associated with later missingness were all present at subsequent time points, we can conclude that the differences found in EB profiles across development are not due to missingness.

**Table S7.** Association between latent profile membership at age 3 and missing status at age 5

|  |  |  |
| --- | --- | --- |
|  | OR (95% CI) | aOR (95% CI) |
| Latent profiles (ref. Low symptoms) |  |  |
| High externalizing and low prosocial | **1.50 (1.16-1.94)** | 1.07 (0.82-1.39) |
| Moderate externalizing | **1.26 (1.09-1.46)** | 1.09 (0.94-1.28) |
| Moderate peer problems | **1.22 (1.02-1.45)** | 1.01 (0.85-1.21) |
| High emotional and conduct | **1.76 (1.33-2.32)** | 1.15 (0.87-1.53) |

aOR – adjusted odds ratio for gender, ethnicity, poverty, monoparental family, maternal education

**Table S8.** Association between latent profile membership at age 5 and missing status at age 7

|  |  |  |
| --- | --- | --- |
|  | OR (95% CI) | aOR (95% CI) |
| Latent profiles (ref. Low symptoms) |  |  |
| **High externalizing and moderate emotional** | **2.23 (1.75-2.85)** | **1.56 (1.20-2.01)** |
| Moderate externalizing | **1.27 (1.07-1.50)** | 1.07 (0.90-1.26) |
| Moderate peer problems | **1.31 (1.05-1.63)** | 1.05 (0.85-1.31) |
| Moderate emotional | **1.27 (1.03-1.56)** | 1.02 (0.82-1.26) |

aOR – adjusted odds ratio for gender, ethnicity, poverty, monoparental family, maternal education

**Table S9.**
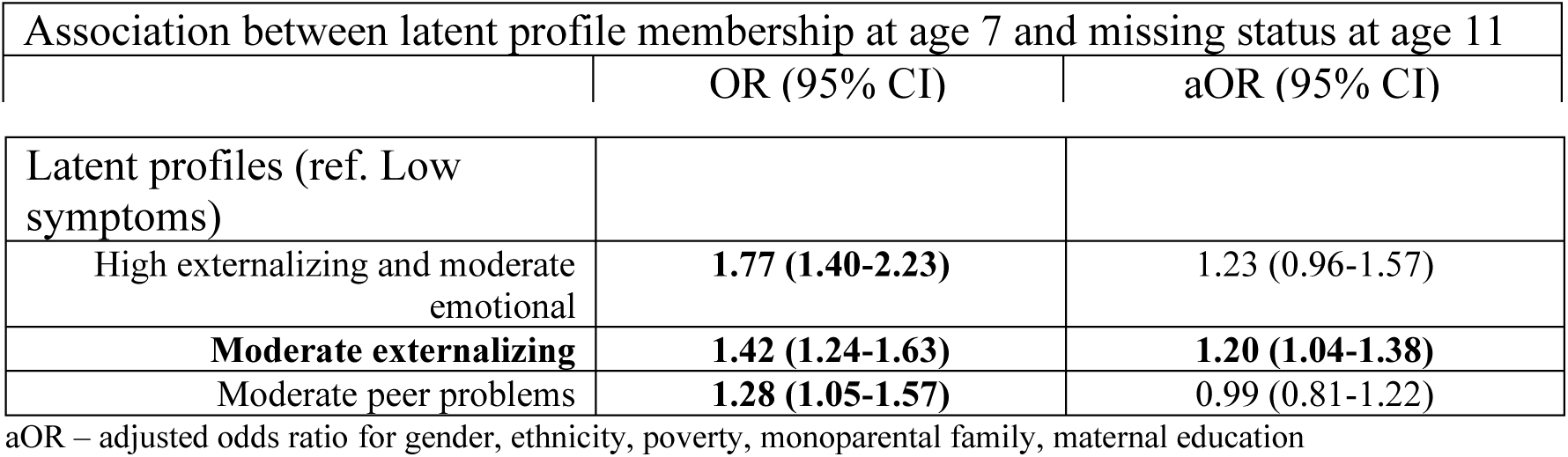
Association between latent profile membership at age 7 and missing status at age 11

|  |  |  |
| --- | --- | --- |
|  | OR (95% CI) | aOR (95% CI) |
| Latent profiles (ref. Low symptoms) |  |  |
| High externalizing and moderate emotional | **1.77 (1.40-2.23)** | 1.23 (0.96-1.57) |
| **Moderate externalizing** | **1.42 (1.24-1.63)** | **1.20 (1.04-1.38)** |
| Moderate peer problems | **1.28 (1.05-1.57)** | 0.99 (0.81-1.22) |

aOR – adjusted odds ratio for gender, ethnicity, poverty, monoparental family, maternal education

**Table S10.** Association between latent profile membership at age 11 and missing status at age 14

|  |  |  |
| --- | --- | --- |
|  | OR (95% CI) | aOR (95% CI) |
| Latent profiles (ref. Low symptoms) |  |  |
| High externalizing and moderate emotional | **1.53 (1.21-1.94)** | 1.16 (0.92-1.48) |
| **Moderate externalizing** | **1.41 (1.22-1.63)** | **1.20 (1.04-1.39)** |
| Moderate internalizing | 1.11 (0.94-1.30) | 0.98 (0.83-1.15) |

aOR – adjusted odds ratio for gender, ethnicity, poverty, monoparental family, maternal education

##### Complete case analysis

We also performed latent profile analyses with complete cases (complete data at all ages). The sample size decreased substantially (n=8083). At age 3, for example, this was a 46% decrease in sample size. The number of profiles, as suggested by the Lo-Mendell-Rubin test, also changed substantially. For ages 3 and 5, the best models have only 3 profiles (previously 5). For age 7 the best model fit remains at 4 profiles. For age 11 the number of profiles increased to 5, and for age 14 decreased from 5 profiles to only 2 profiles. When we extract the same number of profiles as in the original analysis (ignoring the non-significant LMRT p-value), we get similar profiles.

##### Five-profile solutions for ages 7 and 11, and latent profile analysis at time t with complete cases at time t+1

However, as so much information was lost with the complete case analysis, it is difficult to draw any conclusions regarding the effect of missing cases on latent profiles. Thus, we extended the analysis previously performed as follows.

At ages 3, 5, and 14 the LMRT points to 5-profile solutions for these ages. Contrastingly, for ages 7 and 11 the LMRT points to a 4-profile solution. So, one of the major aspects of this analysis is that from age 5 to 7 we have one less profile. The profile that disappears from age 5 to age 7 is the Moderate emotional (6.4%). All the other profiles that are found at age 5 are also found at age 7.

With the regression analyses, we found that membership to the age 5 Moderate emotional profile is not associated with being missing at age 7. Thus, the lack of an age 7 Moderate emotional profile is not due to missing cases from age 5 to age 7.

However, one can argue that if we have chosen a 5-profile solution at age 7 (ignoring the nonsignificant LMRT p-value), the Moderate emotional profile would have appeared.

With this in mind, we extracted a 5-profile solution for age 7:

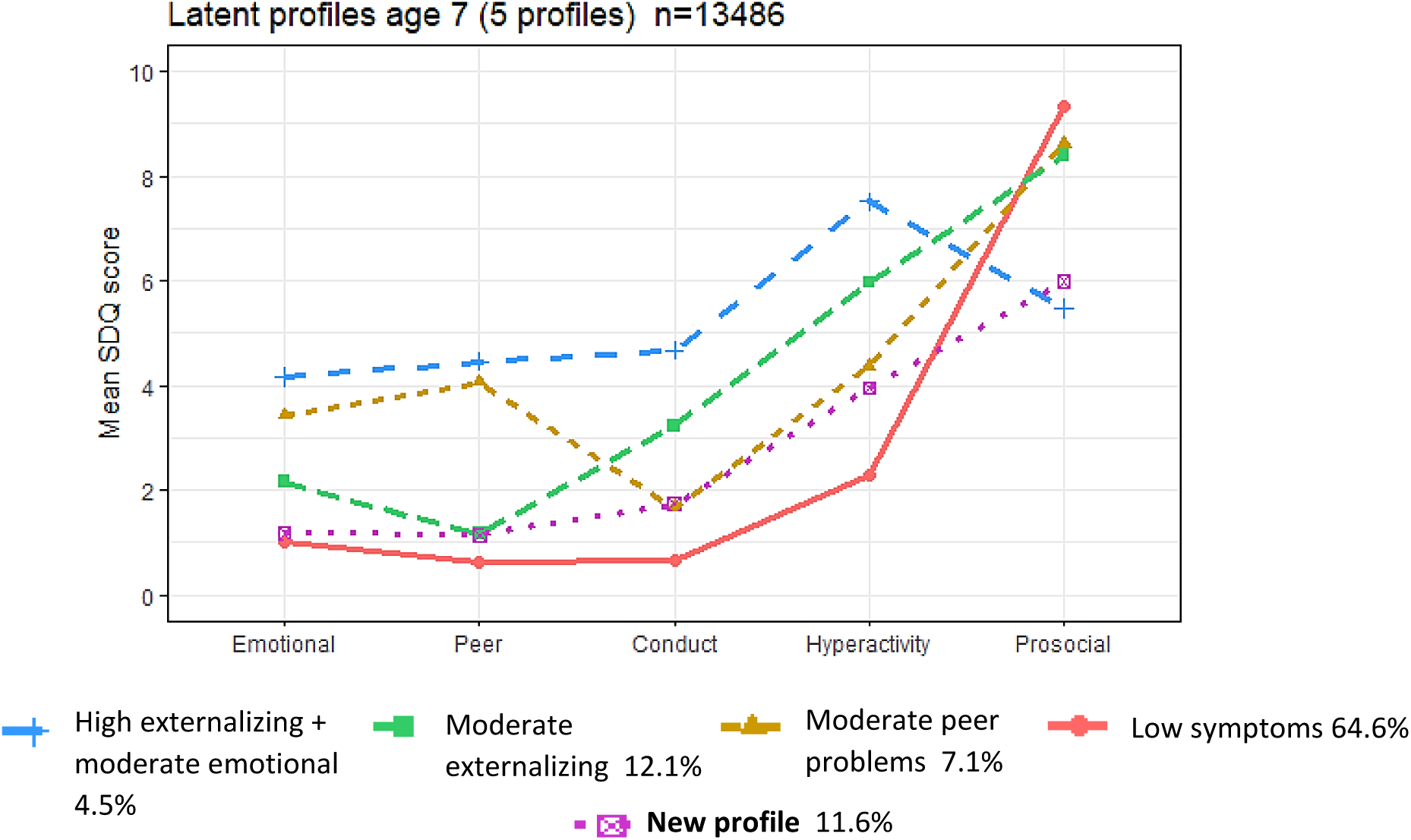

As can be seen in the above figure the New profile (11.6%) that appears in the 5-profile solution is not the Moderate internalizing, but a profile very close to the low symptoms profile, with subthreshold externalizing symptoms. Looking at the class prevalences, this new profile appears to be a subdivision of the Moderate externalizing profile, because comparing to the original 4-profile solution all the prevalences remain similar, except for Moderate externalizing that decreases from 17.9% to 12.1%.

And below is the original 4-profile solution for comparison:

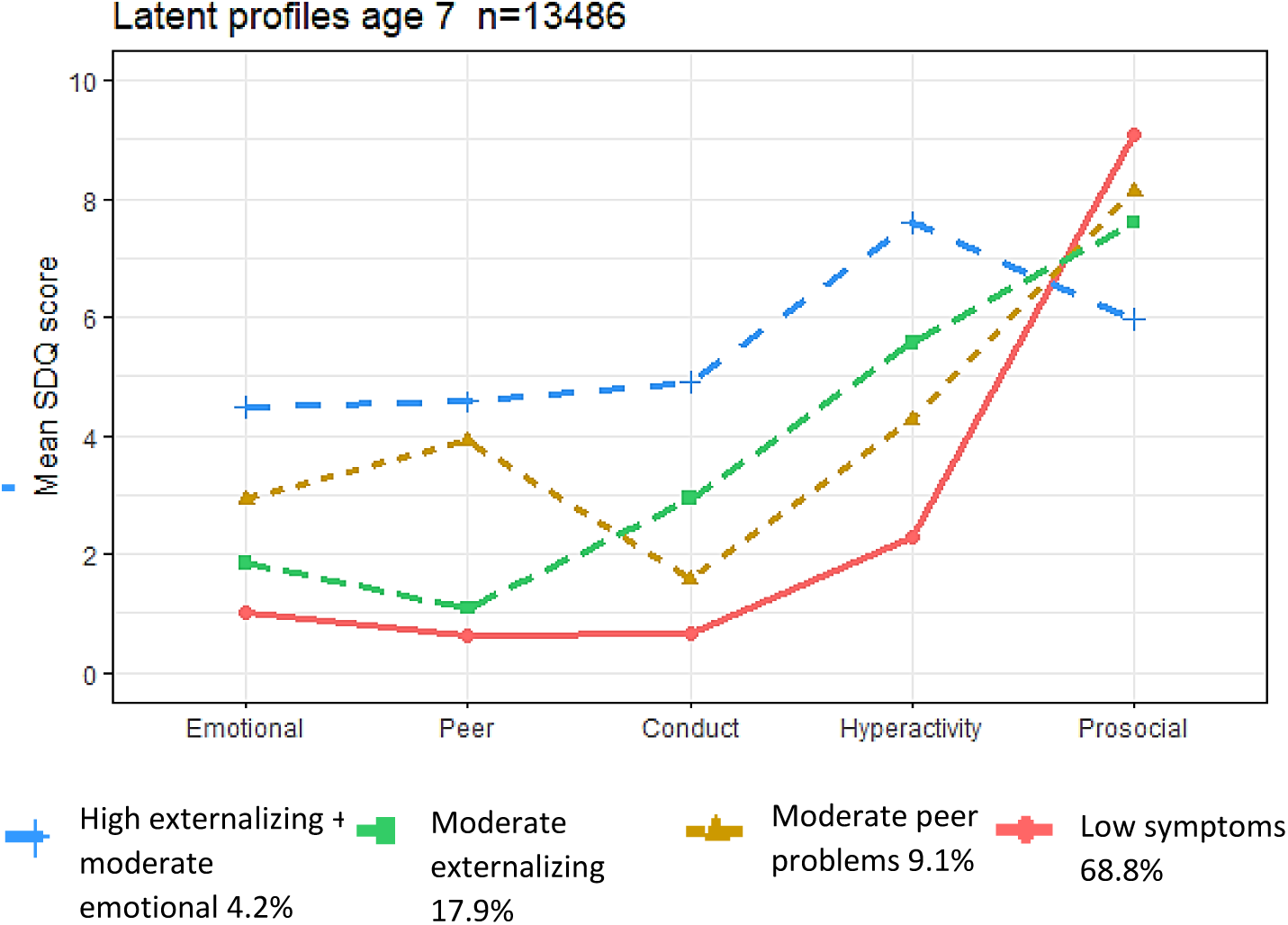

Therefore, even when we try to force a 5-profile solution at age 7 the profiles remain heterogeneous between ages, and the unique profile age 5 Moderate internalizing does not appear at age 7.

If we exclude at age 5 cases that will be missing at age 7, are we able to extract a similar 5-profile solution at age 5?

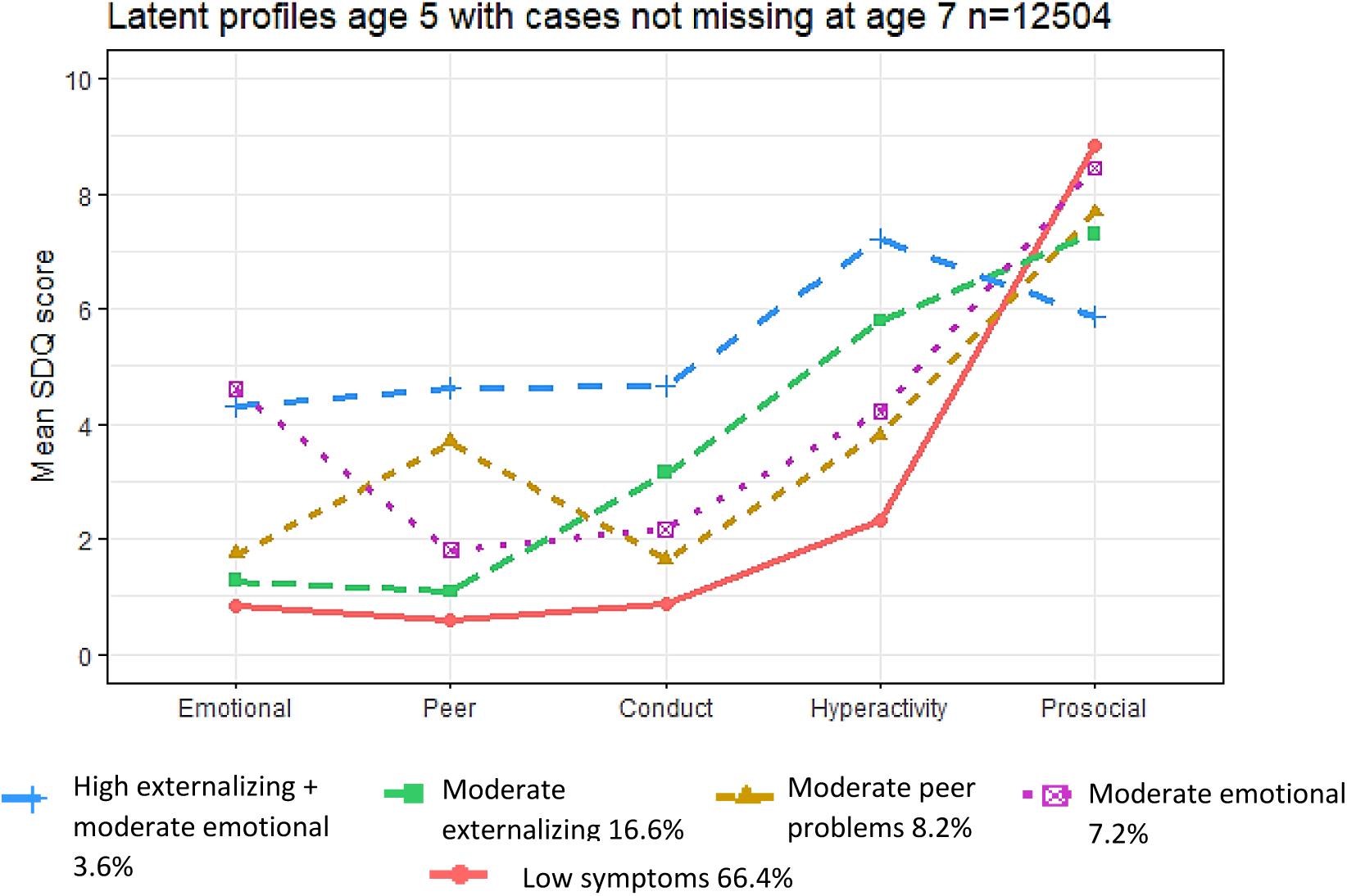

As you can see the profiles are the same as in the original analysis shown below.

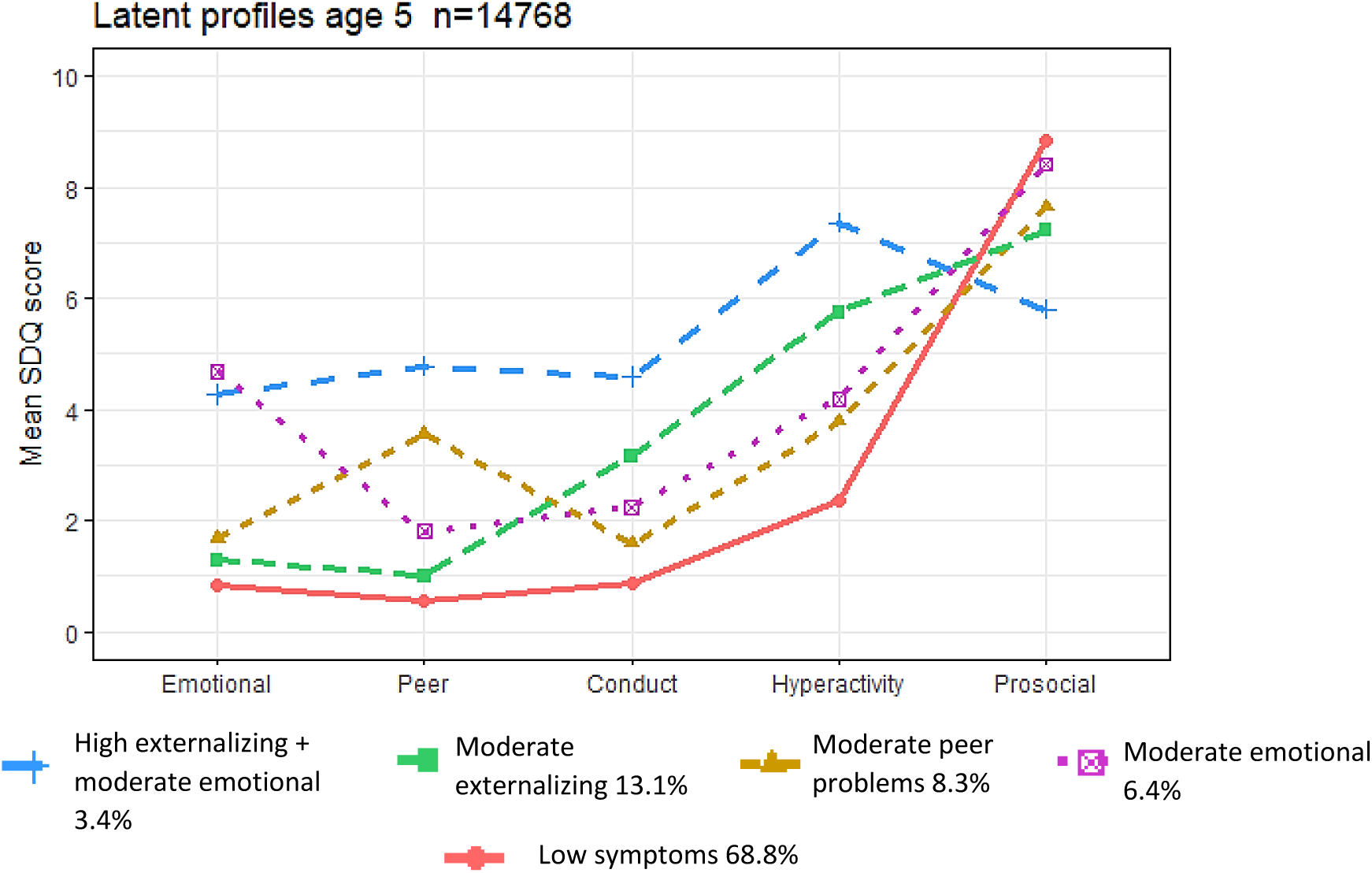

The same rationale applies to the age 11 profiles. The original solution included 4 profiles. If we ignored the non-significant LMRT p-value and forced a 5-profile solution at age 11, would we be able to identify similar profiles to the age 14 5-profile solution?

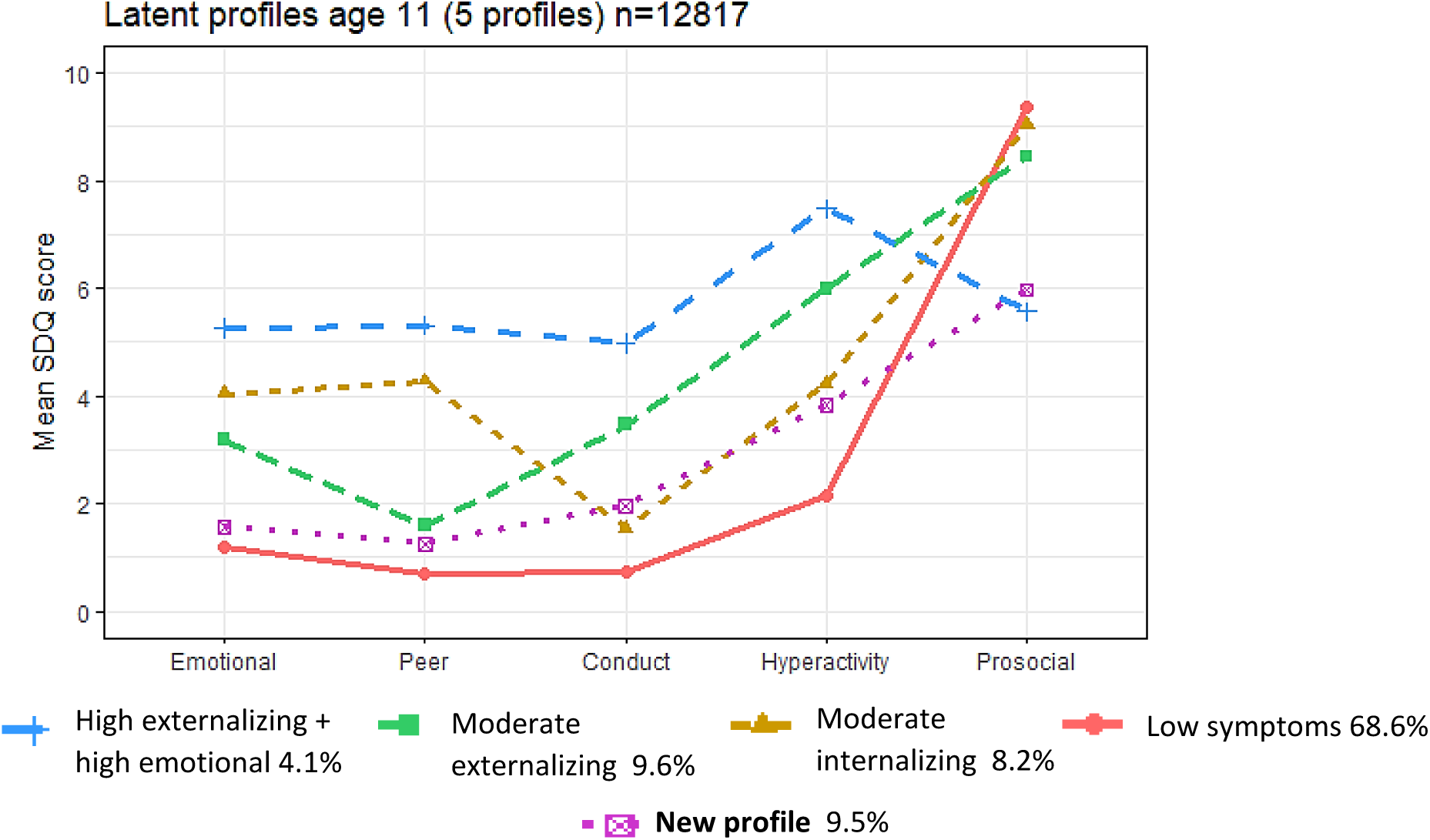

Interestingly, the above figure looks remarkably similar to the age 7 5-profile model, and not with age 14 profiles. Again, a new profile emerges, which is similar to the low symptoms profile, but with slightly higher externalizing symptoms, still subthreshold. Thus, this new profile is not relevant clinically. The similarity between the 5-model profiles between age 7 and age 11, also happens with the 4-profile solutions, as in the original analysis.

This sensitivity analysis, along with the regression analysis of being missing ate time t+1 on membership to latent profiles at time t, support our results: the higher number of profiles for ages 3, 5 and 14 suggests increased heterogeneity in the presentation of emotional and behavioural difficulties when compared to late childhood and preadolescence (ages 7 and 11).

#### S5. Latent transition analysis - Measurement invariance testing

As we found an equal number of profiles between ages 3 and 5, and ages 7 and 11, we estimated three models with partial measurement invariance (a model with equal profiles across ages 3 and 5; a model with equal profiles across ages 7 and 11; and a model with equal profiles across ages 3 and 5, and 7 and 11). Testing for full measurement invariance was not possible because of the different number of profiles found across development. The model with measurement noninvariance had a lower BIC (1,234,036) than models with equal profiles across ages 3 and 5 (BIC=1,268,788); across ages 7 and 11 (BIC = 1,264,576); and across ages 3 and 5, and 7 and 11 (BIC=1,269,297). The likelihood ratio tests indicated that the model with measurement noninvariance provided a better fit to the data, comparing with the partial measurement invariance models. We then chose the model with measurement non-invariance as the best model, which was used in subsequent analyses.

To test if measurement non-invariance would still hold if we have chosen four-profile solutions for ages 3, 5 and 14, we tested for full measurement invariance and measurement non-invariance in LTA models with four-profile solutions across time. The model with measurement noninvariance had a lower BIC (1,268,449) than the model with full measurement invariance (BIC = 1,279,205). The models with partial measurement invariance also had higher BIC than the model with measurement non-invariance.

#### S6. Latent transition probabilities

**Table S11.** Latent transition probabilities (age 3 to age 5)

|  | Age 5 |  |  |  |  |
| --- | --- | --- | --- | --- | --- |
| Age 3 | Low symptoms | Moderate emotional | Moderate peer problems | Moderate externalizing | High externalizing + moderate emotional |
| Low symptoms | 0.932 | 0.022 | 0.037 | 0.008 | 0 |
| High emotional + conduct | 0.145 | 0.384 | 0.134 | 0.163 | 0.175 |
| Moderate peer problems | 0.505 | 0.19 | 0.275 | 0.087 | 0.014 |
| Moderate externalizing | 0.311 | 0.077 | 0.076 | 0.484 | 0.051 |
| High externalizing+ low prosocial | 0.05 | 0.078 | 0.124 | 0.366 | 0.383 |

**Table S12.** Latent transition probabilities (age 5 to age 7)

|  | Age 7 |  |  |  |
| --- | --- | --- | --- | --- |
| Age 5 | Low symptoms | Moderate peer problems | Moderate externalizing | High externalizing+ moderate emotional |
| Low symptoms | 0.909 | 0.036 | 0.055 | 0 |
| Moderate emotional | 0.323 | 0.277 | 0.323 | 0.078 |
| Moderate peer problems | 0.345 | 0.477 | 0.128 | 0.049 |
| Moderate externalizing | 0.122 | 0.036 | 0.743 | 0.099 |
| High externalizing+ moderate emotional | 0 | 0.184 | 0.163 | 0.653 |

**Table S13.** Latent transition probabilities (age 7 to age 11)

| Age 7 | Age 11 |  |  |  |
| --- | --- | --- | --- | --- |
|  | Low symptoms | Moderate internalizing | Moderate externalizing | High externalizing+ high emotional |
| Low symptoms | 0.924 | 0.050 | 0.024 | 0.001 |
| Moderate peer problems | 0.335 | 0.444 | 0.144 | 0.077 |
| Moderate externalizing | 0.264 | 0.097 | 0.580 | 0.059 |
| High externalizing+ moderate emotional | 0.027 | 0.151 | 0.347 | 0.475 |

**Table S14.** Latent transition probabilities (age 11 to age 14)

| Age 11 | Age 14 |  |  |  |  |
| --- | --- | --- | --- | --- | --- |
|  | Low symptoms | High internalizing | Moderate internalizing | Moderate externalizing | High externalizing + moderate emotional |
| Low symptoms | 0.834 | 0.006 | 0.084 | 0.073 | 0.003 |
| Moderate internalizing | 0.274 | 0.189 | 0.357 | 0.173 | 0.006 |
| Moderate externalizing | 0.104 | 0.104 | 0.088 | 0.609 | 0.095 |
| High externalizing + high emotional | 0.003 | 0.332 | 0.037 | 0.245 | 0.384 |

#### S7. Latent transition analysis with covariates

**Table S15.** Latent transition analysis with covariates (age 3)

|  | Age 3 - Odds Ratio (95% CI) \| Reference class = Low symptoms |  |  |  |
| --- | --- | --- | --- | --- |
| Predictors | High externalizing + low prosocial | Moderate externalizing | Moderate peer problems | High emotional + conduct |
|  | OR (95% CI) | OR (95% CI) | OR (95% CI) | OR (95% CI) |
| Gender (male) | **3.01 (2.43-3.74)** | **1.78 (1.58-1.99)** | **1.37 (1.2-1.55)** | 1.15 (0.97-1.36) |
| Ethnicity (white) | **0.48 (0.37-0.64)** | 0.99 (0.81-1.21) | **0.49 (0.41-0.6)** | **0.4 (0.31-0.42)** |
| Monoparental family | **3.68 (2.89-4.68)** | **2.05 (1.75-2.39)** | **1.32 (1.08-1.63)** | **2.81 (2.23-3.54)** |
| Poverty | **5.33 (4.15-6.84)** | **2.67 (2.33-3.07)** | **2.04 (1.75-2.39)** | **5.49 (4.38-6.89)** |
| Maternal professional qualification (high) | **0.26 (0.18-0.37)** | **0.43 (0.37-0.5)** | **0.49 (0.41-0.59)** | **0.24 (0.18-0.34)** |
| Maternal distress | **1.32 (1.29-1.35)** | **1.2 (1.18-1.23)** | **1.15 (1.12-1.18)** | **1.33 (1.3-1.37)** |
| Harsh parenting | **1.41 (1.31-1.52)** | **1.37 (1.33-1.42)** | **1.06 (1.03-1.1)** | **1.21 (1.14-1.29)** |

**Table S16.** Latent transition analysis with covariates (age 5)

|  | Age 5 - Odds Ratio (95% CI) \| Reference class = Low symptoms |  |  |  |
| --- | --- | --- | --- | --- |
| Predictors | High externalizing + moderate emotional | Moderate externalizing | Moderate peer problems | Moderate emotional |
|  | OR (95% CI) | OR (95% CI) | OR (95% CI) | OR (95% CI) |
| Gender (male) | **1.57 (1.27-1.95)** | **1.41 (1.18-1.68)** | **1.29 (1.09-1.52)** | **0.73 (0.61-0.87)** |
| Ethnicity (white) | 0.78 (0.52-1.17) | 1.022 (0.71-1.46) | **0.53 (0.42-0.67)** | **0.58 (0.46-0.75)** |
| Monoparental family | **2.57 (1.96-3.36)** | **1.87 (1.47-2.37)** | **1.72 (1.36-2.09)** | **1.95 (1.53-2.49)** |
| Poverty | **3.5 (2.62-4.68)** | **2.14 (1.67-2.75)** | **1.65 (1.35-2.02)** | **1.92 (1.52-2.42)** |
| Maternal professional qualification (high) | **0.54 (0.38-0.77)** | **0.76 (0.6-0.97)** | **0.77 (0.61-0.96)** | **0.68 (0.52-0.91)** |
| Maternal distress | **1.29 (1.26-1.33)** | **1.15 (1.12-1.18)** | **1.14 (1.11-1.17)** | **1.21 (1.18-1.24)** |
| Harsh parenting | **1.44 (1.34-1.55)** | **1.37 (1.30-1.44)** | **1.08 (1.03-1.13)** | **1.21 (1.15-1.28)** |

**Table S17.** Latent transition analysis with covariates (age 7)

|  | Age 7 - Odds Ratio (95% CI) \| Reference class = Low symptoms |  |  |
| --- | --- | --- | --- |
| Predictors | High externalizing + moderate emotional | Moderate externalizing | Moderate peer problems |
|  | OR (95% CI) | OR (95% CI) | OR (95% CI) |
| Gender (male) | **2.29 (1.75-3.01)** | **1.86 (1.5-2.32)** | 1.18 (0.99-1.42) |
| Ethnicity (white) | 0.91 (0.59-1.41) | 0.91 (0.67-1.24) | **0.54 (0.42-0.69)** |
| Monoparental family | **2.39 (1.68-3.43)** | **1.66 (1.29-2.13)** | **1.76 (1.33-2.34)** |
| Poverty | **2.6 (1.82-3.72)** | **2.07 (1.62-2.63)** | **2.3 (1.82-2.91)** |
| Maternal professional qualification (high) | **0.42 (0.28-0.63)** | **0.73 (0.57-0.93)** | **0.59 (0.47-0.74)** |
| Maternal distress | **1.5 (1.22-1.30)** | **1.14 (1.11-1.18)** | **1.15 (1.12-1.18)** |
| Harsh parenting | **1.75 (1.59-1.93)** | **1.62 (1.52-1.73)** | **1.1 (1.09-1.24)** |

**Table S18.**
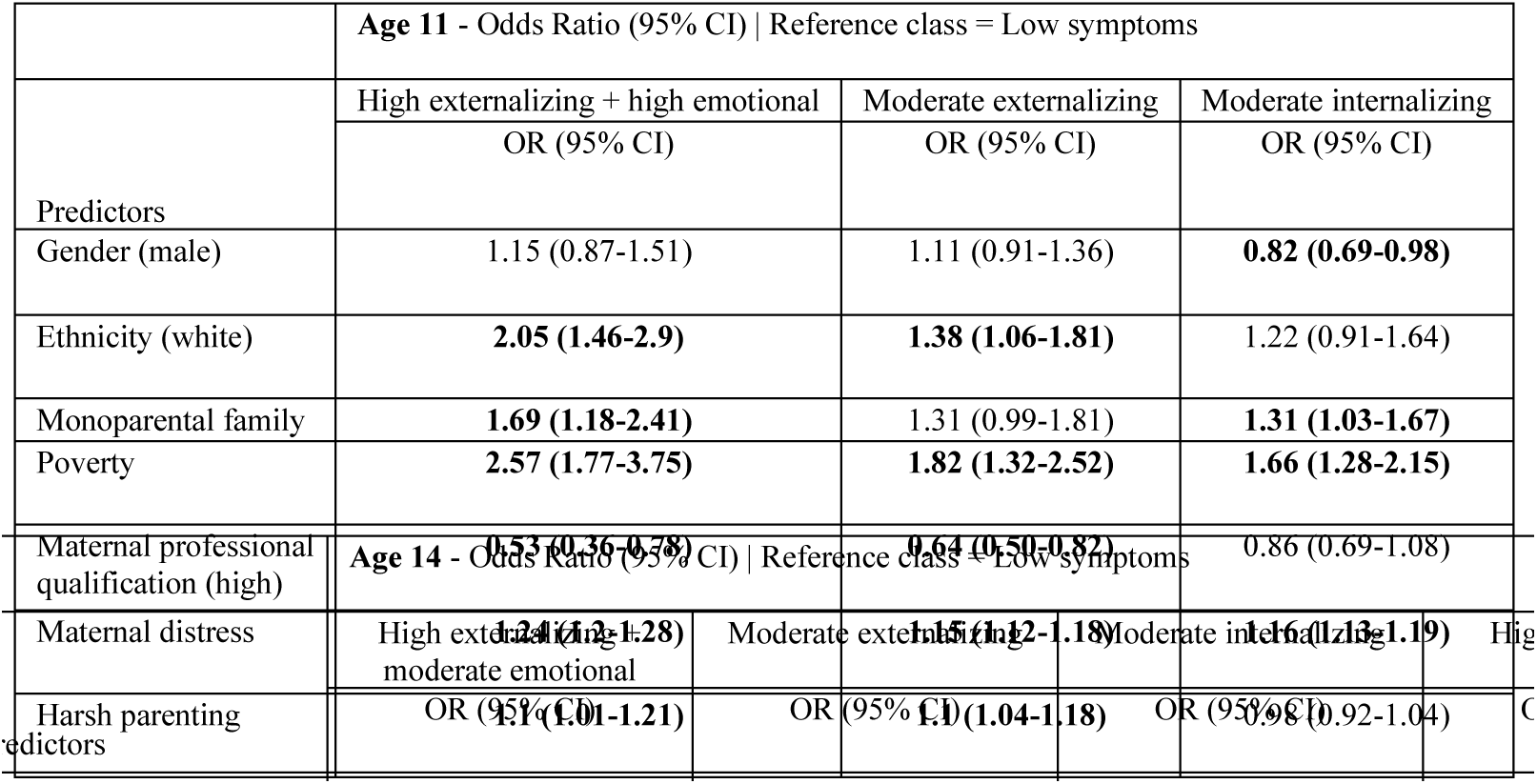
Latent transition analysis with covariates (age 11)

**Table S19.**
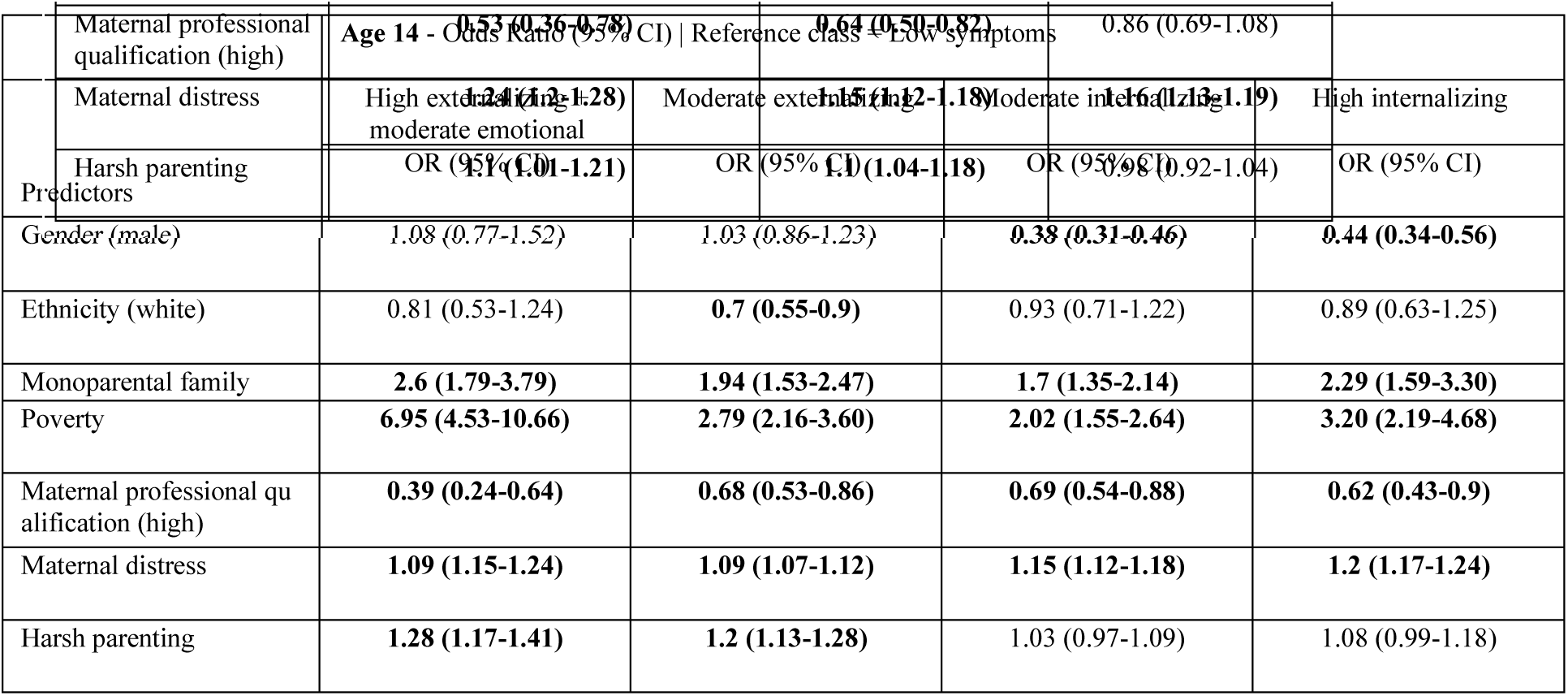
Latent transition analysis with covariates (age 14)

#### S8. Latent transition analysis with covariates influencing transition probabilities

**Table S20.** Covariates influencing transition probabilities

|  | **Age 3 Moderate Externalizing** - Odds Ratio (95% CI) \| Reference class = Age 5 Low symptoms |  |  |
| --- | --- | --- | --- |
| Predictors | To age 5 High externalizing+ moderate emotional | To age 5 Moderate externalizing | To age 5 Moderate emotional |
|  | OR (95% CI) | OR (95% CI) | OR (95% CI) |
| Gender (male) | ns | ns | ns |
| Ethnicity (white) | ns | ns | ns |
| Monoparental family | 2.489 (1.294-4.786) | 1.572 (1.052-2.349) | ns |
| Poverty | 1.787 (1.061-3.010) | ns | ns |
| Maternal professional qualification (high) | 0.440 (0.196-0.987) | ns | ns |
| Maternal distress | 1.099 (1.030-1.173) | ns | 1.112 (1.047-1.181) |
| Harsh parenting | ns | ns | ns |

**Table S21.** Covariates influencing transition probabilities (continued)

|  | **Age 3 Moderate peer problems** - Odds Ratio (95% CI) \| Reference class = Age 5 Low symptoms |  |
| --- | --- | --- |
| Predictors | To age 5 High externalizing+ moderate emotional | To age 5 Moderate emotional |
|  | OR (95% CI) | OR (95% CI) |
| Gender (male) | ns | ns |
| Ethnicity (white) | ns | ns |
| Monoparental family | ns | ns |
| Poverty | ns | 1.736 (1.050-2.872) |
| Maternal professional qualification (high) | ns | ns |
| Maternal distress | ns | 1.072 (1.020-1.127) |
| Harsh parenting | 0.71 (0.5553-0.912) | ns |

**Table S22.** Covariates influencing transition probabilities (continued)

|  | **Age 3 High emotional+conduct** – Odds Ratio (95% CI) \| Reference class = Age 5 Low symptoms |  |  |
| --- | --- | --- | --- |
| Predictors | To age 5 High externalizing+ moderate emotional | To age 5 Moderate emotional | To age 5 Moderate peer problems |
|  | OR (95% CI) | OR (95% CI) | OR (95% CI) |
| Gender (male) | ns | ns | ns |
| Ethnicity (white) | ns | ns | ns |
| Monoparental family | 11.72 (1.103-124.514) |  | ns |
| Poverty | 4.862 (1.839-12.853) | ns | 3.838 (1.160-12.702) |
| Maternal professional qualification (high) | ns | ns | ns |
| Maternal distress | ns | ns | ns |
| Harsh parenting | ns | ns | ns |

**Table S23.** Covariates influencing transition probabilities (continued)

|  | **Age 3 Low symptoms** - Odds Ratio (95% CI) \| Reference class = Age 5 Low symptoms |  |
| --- | --- | --- |
| Predictors | To age 5 Moderate peer problems | To age 5 Moderate emotional |
|  | OR (95% CI) | OR (95% CI) |
| Gender (male) | ns | ns |
| Ethnicity (white) | ns | ns |
| Monoparental family | 2.251 (1.373-3.690) | 3.424 (1.711-6.854) |
| Poverty | 3.476 (2.175-5.558) | 2.994 (1.508-5.945) |
| Maternal professional qualification (high) | ns | ns |
| Maternal distress | ns | 1.113 (1.025-1.210) |
| Harsh parenting | 0.88 (0.777-0.996) | ns |

**Table S24.** Covariates influencing transition probabilities (continued)

|  | **Age 5 Moderate externalizing** - Odds Ratio (95% CI) \| Reference class = Age 7 Low symptoms |  |  |
| --- | --- | --- | --- |
| Predictors | To age 7 High externalizing+ moderate emotional | To age 7 Moderate externalizing | To age 7 Moderate peer problems |
|  | OR (95% CI) | OR (95% CI) | OR (95% CI) |
| Gender (male) | ns | ns | ns |
| Ethnicity (white) | ns | ns | ns |
| Monoparental family | 2.992 (1.320-6.784) | ns | 4.881 (1.241-19.191) |
| Poverty | 2.528 (1.320-4.842) | ns | ns |
| Maternal professional qualification (high) | ns | ns | ns |
| Maternal distress | ns | ns | ns |
| Harsh parenting | 1.234 (1.039-1.466) | 1.134 (1.001-1.284) | ns |

**Table S25.** Covariates influencing transition probabilities (continued)

|  | **Age 5 Moderate peer problems** - Odds Ratio (95% CI) \| Reference class = Age 7 Low symptoms |  |
| --- | --- | --- |
| Predictors | To age 7 High externalizing+ moderate emotional | To age 7 Moderate peer problems |
|  | OR (95% CI) | OR (95% CI) |
| Gender (male) | ns | ns |
| Ethnicity (white) | ns | ns |
| Monoparental family | ns | ns |
| Poverty | ns | ns |
| Maternal professional qualification (high) | ns | ns |
| Maternal distress | ns | 1.10 (1.037-1.188) |
| Harsh parenting | 1.409 (1.146-1.733) | 1.163 (1.037-1.303) |

**Table S26.** Covariates influencing transition probabilities (continued)

|  | **Age 5 Moderate emotional** - Odds Ratio (95% CI) \| Reference class = Age 7 Low symptoms |  |  |
| --- | --- | --- | --- |
| Predictors | To age 7 High externalizing+<br>moderate emotional | To age 7 Moderate<br>externalizing | To age 7 Moderate<br>peer problems |
|  | OR (95% CI) | OR (95% CI) | OR (95% CI) |
| Gender (male) | ns | ns | ns |
| Ethnicity (white) | ns | ns | ns |
| Monoparental family | ns | ns | ns |
| Poverty | ns | ns | ns |
| Maternal professional<br>qualification (high) | ns | ns | ns |
| Maternal distress | 1.118 (1.026-1.218) | ns | 1.077 (1.005-1.155) |
| Harsh parenting | ns | 1.218 (1.062-1.398) | ns |

**Table S27.** Covariates influencing transition probabilities (continued)

|  | **Age 5 Low symptoms** - Odds Ratio (95% CI) \| Reference class = Age 7 Low symptoms |  |
| --- | --- | --- |
| Predictors | To age 7 Moderate<br>externalizing | To age 7 Moderate<br>peer problems |
|  | OR (95% CI) | OR (95% CI) |
| Gender (male) | ns | ns |
| Ethnicity (white) | ns | ns |
| Monoparental family | 1.823 (1.199-2.771) | ns |
| Poverty | 2.671 (1.786-3.993) | 3.049 (1.964-4.734) |
| Maternal professional<br>qualification (high) | 0.596 (0.398-0.894) | 0.446 (0.273-0.730) |
| Maternal distress | 1.134 (1.088-1.181) | 1.112 (1.061-1.165) |
| Harsh parenting | 1.358 (1.205-1.531) | ns |

**Table S28.** Covariates influencing transition probabilities (continued)

|  | **Age 7 High externalizing + moderate emotional** - Odds Ratio (95% CI) \|<br>Reference class = Age 11 Low symptoms |  |  |
| --- | --- | --- | --- |
| Predictors | To age 11 High externalizing+<br>high emotional | To age 11 Moderate<br>externalizing | To age 11 Moderate<br>internalizing |
|  | OR (95% CI) | OR (95% CI) | OR (95% CI) |
| Gender (male) | ns | ns | ns |
| Ethnicity (white) | ns | ns | ns |
| Monoparental family | ns | ns | ns |
| Poverty | ns | ns | ns |
| Maternal professional<br>qualification (high) | ns | ns | ns |
| Maternal distress | ns | ns | ns |
| Harsh parenting | 1.934 (1.028-3.638) | 1.939 (1.041-3.609) | ns |

**Table S29.** Covariates influencing transition probabilities (continued)

|  | **Age 7 Moderate peer problems** - Odds Ratio (95% CI) \| Reference class = Age 11 Low symptoms |
| --- | --- |
|  | To age 11 High externalizing+high emotional<br>OR (95% CI) |
| Predictors |  |
| Gender (male) | ns |
| Ethnicity (white) | ns |
| Monoparental family | ns |
| Poverty | ns |
| Maternal professional qualification (high) | ns |
| Maternal distress | 1.113 (1.042-1.190) |
| Harsh parenting | 1.24 (1.036-1.483) |

**Table S30.**
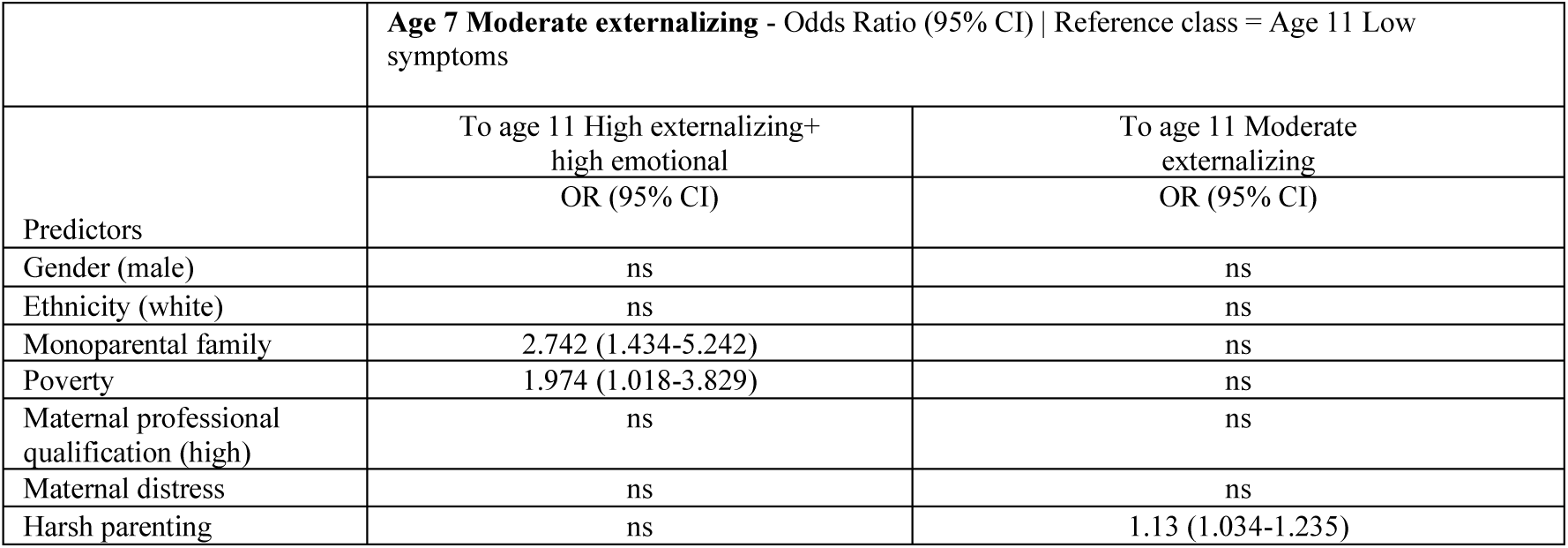
Covariates influencing transition probabilities (continued)

|  | **Age 7 Moderate externalizing** - Odds Ratio (95% CI) \| Reference class = Age 11 Low symptoms |  |
| --- | --- | --- |
|  | To age 11 High externalizing+<br>high emotional<br>OR (95% CI) | To age 11 Moderate<br>externalizing<br>OR (95% CI) |
| Predictors |  |  |
| Gender (male) | ns | ns |
| Ethnicity (white) | ns | ns |
| Monoparental family | 2.742 (1.434-5.242) | ns |
| Poverty | 1.974 (1.018-3.829) | ns |
| Maternal professional qualification (high) | ns | ns |
| Maternal distress | ns | ns |
| Harsh parenting | ns | 1.13 (1.034-1.235) |

**Table S31.**
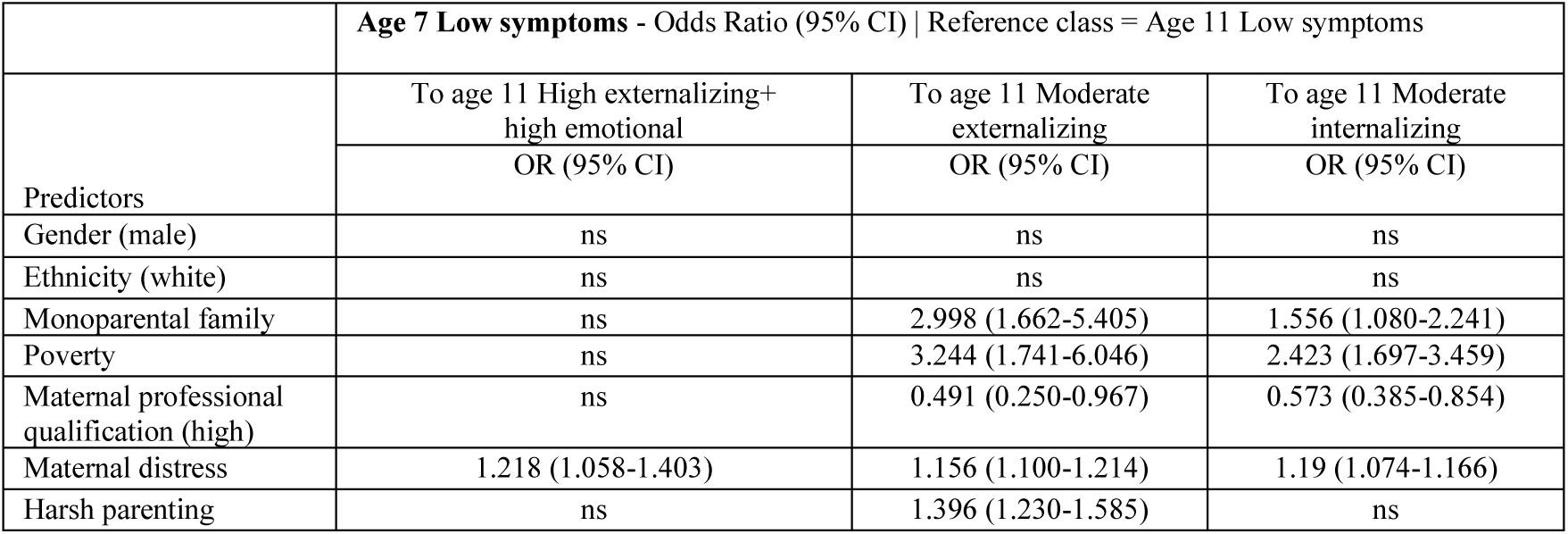
Covariates influencing transition probabilities (continued)

**Table S32.** Covariates influencing transition probabilities (continued)

|  | **Age 11 Moderate internalizing** - Odds Ratio (95% CI) \| Reference class = Age 14 Low symptoms |  |  |
| --- | --- | --- | --- |
| Predictors | To age 14 High internalizing | To age 14 Moderate externalizing | To age 14 Moderate internalizing |
|  | OR (95% CI) | OR (95% CI) | OR (95% CI) |
| Gender (male) | ns | ns | ns |
| Ethnicity (white) | ns | ns | ns |
| Monoparental family | ns | ns | ns |
| Poverty | 2.446 (1.273-4.702) | ns | ns |
| Maternal professional qualification (high) | 0.506 (0.264-0.971) | ns | ns |
| Maternal distress | 1.132 (1.067-1.201) | ns | 1.075 (1.002-1.153) |
| Harsh parenting | ns | 1.257 (1.024-1.542) | ns |

**Table S33.**
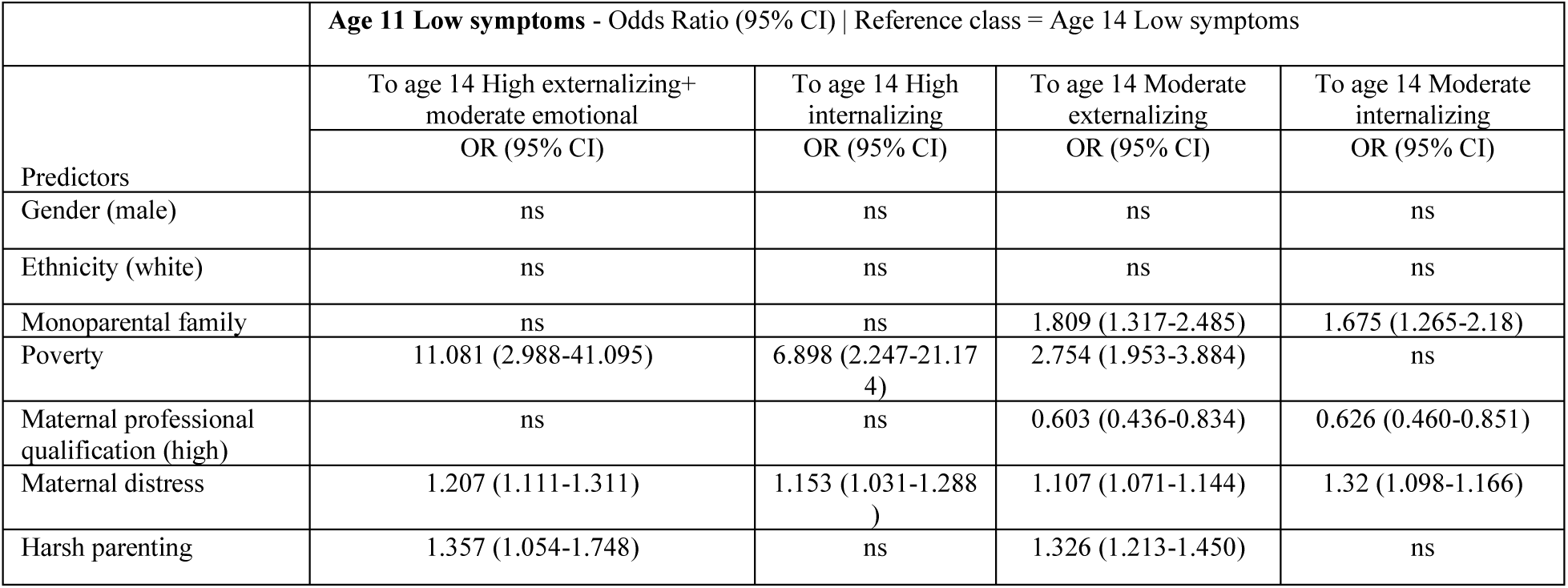
Covariates influencing transition probabilities (continued)

|  | **Age 11 Low symptoms** - Odds Ratio (95% CI) \| Reference class = Age 14 Low symptoms |  |  |  |
| --- | --- | --- | --- | --- |
| Predictors | To age 14 High externalizing+ moderate emotional | To age 14 High internalizing | To age 14 Moderate externalizing | To age 14 Moderate internalizing |
|  | OR (95% CI) | OR (95% CI) | OR (95% CI) | OR (95% CI) |
| Gender (male) | ns | ns | ns | ns |
| Ethnicity (white) | ns | ns | ns | ns |
| Monoparental family | ns | ns | 1.809 (1.317-2.485) | 1.675 (1.265-2.18) |
| Poverty | 11.081 (2.988-41.095) | 6.898 (2.247-21.174) | 2.754 (1.953-3.884) | ns |
| Maternal professional qualification (high) | ns | ns | 0.603 (0.436-0.834) | 0.626 (0.460-0.851) |
| Maternal distress | 1.207 (1.111-1.311) | 1.153 (1.031-1.288) | 1.107 (1.071-1.144) | 1.32 (1.098-1.166) |
| Harsh parenting | 1.357 (1.054-1.748) | ns | 1.326 (1.213-1.450) | ns |

#### S9. Mplus syntax and R code

##### A. Latent profile analysis (Figure 1)

*We repeated this section of the analysis for each age (3, 5, 7, 11, 14)*.

TITLE: Latent profile analysis age 14;

DATA:

FILE IS *“the directory of the dataset in .dat format”;*

VARIABLE:

NAMES = ID PTTYPE2 SPTN00 WEIGHT2 emo_3 cond_3 hype_3

> peers_3 prosoc_3 emo_5 cond_5 hype_5 peers_5 prosoc_5 emo_7 cond_7 hype_7 peers_7 prosoc_7 emo_11 cond_11 hype_11 peer_11 prosoc_11 emo_14 cond_14 hyper_14 peer_14 prosoc_14;

MISSING=.;

IDVARIABLE IS ID;

WEIGHT IS weight2;

STRATIFICATION IS pttype2;

CLUSTER IS sptn00;

usevar= EMO_14 COND_14 HYPER_14 PEER_14 PROSOC_14; *!emotional-behavioral SDQ !subscales for age 14*

CLASSES = c (5); *!test different number of classes, changing the number in brackets*

ANALYSIS:

TYPE IS MIXTURE COMPLEX; *!type is complex when the user specifies weight, stratification and !cluster variables*

STARTS = 2000 500; *!raise number of starting values to improve identification*

LRTSTARTS =0 0 200 50; *!starts for the Lo-Mendell-Rubin test; this part of syntax is only !needed if asking in the output for TECH11 or TECH14*

PROCESSORS = 12; !number of CPU cores for parallel computation

PLOT:

type is plot3;

series is EMO_14 (1) COND_14 (2) HYPER_14 (3) PEER_14 (4) PROSOC_14 (5);

OUTPUT: TECH1 TECH11 svalues; *!ask for svalues to reorder the number of profiles if needed; usually, the last profile will be the reference profile*

savedata: !save modal class assignment for each participant, to use in the next step of the analysis file is 3_4clca.dat; !give a name to the new dataset

save = **cprob**; !save modal class assignment

##### B. Latent profile analysis – three-step specification step

This section uses the modal class assignment, and the logits for the classification probabilities for the most likely latent class/profile membership obtained in the previous step, in order to perform the manual three-step specification.

Below is the part of the Mplus output obtained in the previous step with the logits, for the age 14 emotional and behavioral profiles:

Logits for the Classification Probabilities for the Most Likely Latent Class Membership (Column) by Latent Class (Row)

|  | 1 | 2 | 3 | 4 | 5 |
| --- | --- | --- | --- | --- | --- |
| 1 | 9.107 | 6.174 | -4.411 | 5.994 | 0.000 |
| 2 | -2.940 | 1.869 | -1.375 | -2.125 | 0.000 |
| 3 | -10.333 | -1.096 | 1.531 | -1.757 | 0.000 |
| 4 | 5.145 | 6.057 | 6.341 | 8.834 | 0.000 |
| 5 | -13.774 | -3.687 | -4.036 | -12.505 | 0.000 |

And now the Mplus syntax below:

NAMES = ID PTTYPE2 SPTN00 WEIGHT2 emo_3 cond_3 hype_3

> peers_3 prosoc_3 emo_5 cond_5 hype_5 peers_5 prosoc_5 emo_7 cond_7 hype_7
>
> peers_7 prosoc_7 emo_11 cond_11 hype_11 peer_11 prosoc_11 emo_14 cond_14
>
> hyper_14 peer_14 prosoc_14 **cl_14y5**; *! cl_14y5 is the modal class assignment obtained in the previous step with save = cprob, with values of 1 to 5 (each representing a specific latent profile;*

MISSING=.;

IDVARIABLE IS ID;

WEIGHT IS weight2;

STRATIFICATION IS pttype2;

CLUSTER IS sptn00;

NOMINAL= **cl_14y5**;

USEVAR= **cl_14y5**;

CLASSES = C (5);

ANALYSIS:

TYPE IS MIXTURE COMPLEX;

STARTS = 0;

PROCESSORS = 12;

MODEL:

%c#1%

[cl_14y5#1@9.107]; *!notice the logits after the “@” obtained from the output excerpt shown !above*

[cl_14y5#2@6.174];

[cl_14y5#3@-4.411];

[cl_14y5#4@5.994];

%c#2%

[cl_14y5#1@-2.940];

[cl_14y5#2@1.869];

[cl_14y5#3@-1.375];

[cl_14y5#4@-2.125];

%c#3%

[cl_14y5#1@-10.333];

[cl_14y5#2@-1.096];

[cl_14y5#3@1.531];

[cl_14y5#4@-1.757];

%c#4%

[cl_14y5#1@5.145];

[cl_14y5#2@6.057];

[cl_14y5#3@6.341];

[cl_14y5#4@8.834];

%c#5%

[cl_14y5#1@-13.774];

[cl_14y5#2@-3.687];

[cl_14y5#3@-4.036];

[cl_14y5#4@-12.505];

OUTPUT: TECH1 TECH11 sampstat;

Repeat this step for each age (3, 5, 7, 11, and 14). The “MODEL” section of the syntax will be included in the LTA analysis.

##### C. Latent transition analysis with the manual three-step specification, with measurement non-invariance (Figure 2 and tables S11 to S14)

NAMES = ID PTTYPE2 SPTN00 WEIGHT2 emo_3 cond_3 hype_3 peers_3 prosoc_3 emo_5 cond_5 hype_5 peers_5 prosoc_5 emo_7 cond_7 hype_7 peers_7 prosoc_7 emo_11 cond_11 hype_11 peer_11 prosoc_11 emo_14 cond_14 hyper_14 peer_14 prosoc_14 male white harsh_3 harsh_5 harsh_7 pov_9m pov_3 pov_5 pov_7 pov_11 pov_14 high_pro kess_3 kess_5 kess_7 kess_11 kess_14 mono_9m mono_3 mono_5 mono_7 mono_11 mono_14 alc_14 smok_14 binge_14 cann_14 sharm mentw_14 cl_3y5 cl_5y5 cl_3y5 cl_7y cl_11y cl_14y5;

MISSING=.;

IDVARIABLE IS ID;

WEIGHT IS weight2;

STRATIFICATION IS pttype2;

CLUSTER IS sptn00;

nominal= cl_3y5 cl_5y5 cl_7y cl_11y cl_14y5;

usevar= cl_3y5 cl_5y5 cl_7y cl_11y cl_14y5;

CLASSES = C1(5) C2(5) C3(4) C4(4) C5(5); *!C1 is age 3 latent profiles; C2 is age 5; C3 is age !7; C4 is !age 11; C5 is age 14; notice the number of profiles in brackets*

ANALYSIS:

TYPE IS MIXTURE COMPLEX;

STARTS = 0;

PROCESSORS = 12;

MODEL:

%OVERALL%

C2 on C1;

C3 on C2;

C4 on C3;

C5 on C4;

MODEL C1:

%c1#1% *!age 3, latent profile 1*

[cl_3y5#1@8.900];

[cl_3y5#2@6.397];

[cl_3y5#3@6.690];

[cl_3y5#4@5.822];

%c1#2%

[cl_3y5#1@-3.343];

[cl_3y5#2@1.365];

[cl_3y5#3@-1.358];

[cl_3y5#4@-2.970];

%c1#3%

[cl_3y5#1@-2.045];

[cl_3y5#2@-0.313];

[cl_3y5#3@1.928];

[cl_3y5#4@-2.095];

%c1#4%

[cl_3y5#1@0.944];

[cl_3y5#2@1.818];

[cl_3y5#3@1.434];

[cl_3y5#4@4.157];

%c1#5%

[cl_3y5#1@-13.102];

[cl_3y5#2@-2.889];

[cl_3y5#3 @-3.726];

[cl_3y5#4@-7.500];

MODEL C2:

%c2#1%

[cl_5y5#1@9.749];

[cl_5y5#2@5.904];

[cl_5y5#3@6.469];

[cl_5y5#4 @6.243];

%c2#2%

[cl_5y5#1@-3.534];

[cl_5y5#2@1.446];

[cl_5y5#3@-1.833];

[cl_5y5#4@-1.941];

%c2#3%

[cl_5y5#1 @-1.846];

[cl_5y5#2@-G.702];

[cl_5y5#3@2.062];

[cl_5y5#4@-0.993];

%c2#4%

[cl_5y5# 1 @-1.658];

[cl_5y5#2@-0.459];

[cl_5y5#3@-0.636];

[cl_5y5#4 @2.256];

%c2#5%

[cl_5y5#1@-12.651];

[cl_5y5#2@-3.724];

[cl_5y5#3@-4.641];

[cl_5y5#4@-5.127];

MODEL C3:

%C3#1%

[cl_7y#1@13.G59];

[cl_7y#2@10.139];

[cl_7y#3@10.118];

%C3#2%

[cl_7y#1@-2.691];

[cl_7y#2@1.766];

[cl_7y#3 @-1.356];

%C3#3%

[cl_7y# 1 @-1.510];

[cl_7y#2@-0.180];

[cl_7y#3@2.249];

%C3#4%

[cl_7y#1@-13.777];

[cl_7y#2@-3.495];

[cl_7y#3@-4.678];

MODEL C4:

%C4#1%

[cl_11y#1@12.443];

[cl_11y#2@9.568];

[cl_11y#3@9.422];

%C4#2%

[cl_11y#1@-2.702];

[cl_11y#2@1.786];

[cl_11y#3 @-1.287];

%C4#3%

[cl_11y#1@-2.076];

[cl_11y#2@-0.497];

[cl_11y#3@2.156];

%C4#4%

[cl_11y#1@-12.520];

[cl_11y#2@-4.009];

[cl_11y#3@-4.632];

MODEL C5:

%c5#1%

[cl_14y5#1@9.107];

[cl_14y5#2@6.174];

[cl_14y5#3@-4.411];

[cl_14y5#4@5.994];

%c5#2%

[cl_14y5#1@-2.940];

[cl_14y5#2@1.869];

[cl_14y5#3@-1.375];

[cl_14y5#4@-2.125];

%c5#3%

[cl_14y5#1@-10.333];

[cl_14y5#2@-1.096];

[cl_14y5#3@1.531];

[cl_14y5#4@-1.757];

%c5#4%

[cl_14y5#1@5.145];

[cl_14y5#2@6.057];

[cl_14y5#3@6.341];

[cl_14y5#4@8.834];

%c5#5%

[cl_14y5#1@-13.774];

[cl_14y5#2@-3.687];

[cl_14y5#3@-4.036]

[cl_14y5#4@-12.505];

OUTPUT: PATTERNS CINTERVAL RESIDUAL TECH1 TECH8;

From this syntax we obtain the latent transition probabilities:

LATENT TRANSITION PROBABILITIES BASED ON THE ESTIMATED MODEL

C1 Classes (Rows) by C2 Classes (Columns)

|  | 1 | 2 | 3 | 4 | 5 |
| --- | --- | --- | --- | --- | --- |
| 1 | 0.383 | 0.366 | 0.124 | 0.078 | 0.050 |
| 2 | 0.051 | 0.484 | 0.076 | 0.077 | 0.311 |
| 3 | 0.014 | 0.087 | 0.275 | 0.119 | 0.505 |
| 4 | 0.175 | 0.163 | 0.134 | 0.384 | 0.145 |
| 5 | 0.000 | 0.008 | 0.037 | 0.022 | 0.932 |

C2 Classes (Rows) by C3 Classes (Columns)

|  | 1 | 2 | 3 | 4 |
| --- | --- | --- | --- | --- |
| 1 | 0.653 | 0.163 | 0.184 | 0.000 |
| 2 | 0.099 | 0.743 | 0.036 | 0.122 |
| 3 | 0.049 | 0.128 | 0.477 | 0.345 |
| 4 | 0.078 | 0.323 | 0.277 | 0.323 |
| 5 | 0.000 | 0.055 | 0.036 | 0.909 |

C3 Classes (Rows) by C4 Classes (Columns)

|  | 1 | 2 | 3 | 4 |
| --- | --- | --- | --- | --- |
| 1 | 0.475 | 0.347 | 0.151 | 0.027 |
| 2 | 0.059 | 0.580 | 0.097 | 0.264 |
| 3 | 0.077 | 0.144 | 0.444 | 0.335 |
| 4 | 0.001 | 0.024 | 0.050 | 0.924 |

C4 Classes (Rows) by C5 Classes (Columns)

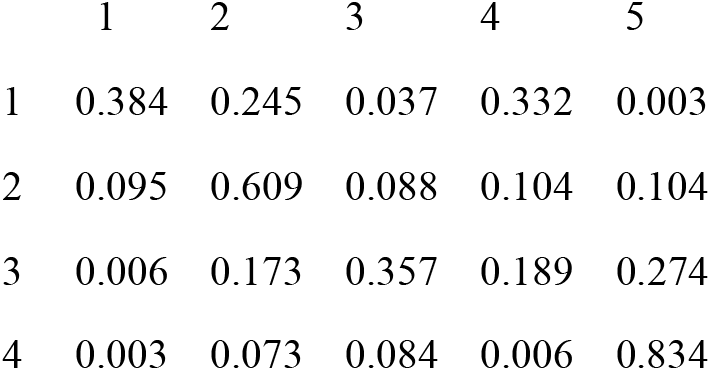

##### D. Latent transition analysis with covariates (Figure 3 and tables S15 to S19)

This part of the syntax builds on the previous section, including covariates in the model. We included each covariate separately, adjusting for gender and ethnicity. Below is the syntax for the covariate maternal psychosocial distress, which was collected at each sweep (time-varying covariate).

For these analyses, we used 25 multiply imputed datasets by chained equations. One can use Mplus built-in capabilities for multiple imputation or other preferred software.

MODEL:

%OVERALL%

C2 on C1;

C3 on C2;

C4 on C3;

C5 on C4;

C1 on male KESS_3 white; !notice that we used here time-varying covariates

C2 on male KESS_5 white;

C3 on male KESS_7 white;

C4 on male KESS_11 white;

C5 on male KESS_14 white;

The rest of the code is the same as in the previous step. When using imputed datasets, one has to specify DATA: TYPE = IMPUTATION.

An excerpt of the confidence interval output obtained from this analysis:

CONFIDENCE INTERVALS FOR THE LOGISTIC REGRESSION ODDS RATIO RESULTS

Categorical Latent Variables

C2#1 ON !C2#1 refers to the age 5 *High externalizing and moderate emotional* profile

|  |  |  |  |  |  |  |  |
| --- | --- | --- | --- | --- | --- | --- | --- |
| MALE | 1.251 | 1.357 | 1.416 | 1.762 | 2.193 | 2.286 | 2.482 |
| KESS_5 | 1.244 | **1.256** | 1.262 | <u>1.294</u> | 1.327 | **1.334** | 1.346 |
| WHITE | 0.491 | 0.571 | 0.617 | 0.923 | 1.382 | 1.493 | 1.736 |

Notice that the middle value is the estimate (underscored), and the bold values are the 95% confidence intervals. Immediately adjacent to the estimates are the 90% confidence intervals. (In the original output these values are not highlighted.)

Then we repeated the analyses for each of the covariates, adjusting for gender and ethnicity.

##### E. Influence of covariates on the transition probabilities (Tables S20 to S33)

In this part of the analysis, we explored the influence of covariates on how the children transition between emotional and behavioural profiles across development. This was achieved allowing for the covariates to have an interaction on the transition probabilities.

This part of the analysis was repeated for each covariate. Below is the code for high maternal professional qualification.

MODEL:

%OVERALL%

C2 on C1

C3 on C2

C4 on C3;

C5 on C4;

C1 on high_pro;

MODEL C1:

%c1#1%

[cl_3y5#1@8.900];

[cl_3y5#2 @6.397];

[cl_3y5#3 @6.690];

[cl_3y5#4@5. 822];

C2 on high_pro; *!the covariate enters here. Notice that in MODEL C1 for age 3 one inserts C2 corresponding to the next sweep (age 5)*.

%c1#2%

[cl_3y5#1@-3.343];

[cl_3y5#2@1.365];

[cl_3y5#3 @-1.358];

[cl_3y5#4@-2.970];

C2 on high_pro;

%c1#3%

[cl_3y5#1@-2.045];

[cl_3y5#2@-0.313];

[cl_3y5#3@1.928];

[cl_3y5#4@-2.095];

C2 on high_pro;

%c1#4%

[cl_3y5#1@0.944];

[cl_3y5#2@1.818];

[cl_3y5#3@1.434];

[cl_3y5#4 @4.157];

C2 on high_pro;

%c1#5%

[cl_3y5#1@-13.102];

[cl_3y5#2@-2.889];

[cl_3y5#3 @-3.726];

[cl_3y5#4@-7.500];

C2 on high_pro;

MODEL C2:

%c2#1%

[cl_5y5#1@9.749];

[cl_5y5#2@5.904];

[cl_5y5#3 @6.469];

[cl_5y5#4 @6.243];

C3 on high_pro; *! Notice that now it is C3 (and not C2 as in MODEL C1)*

%c2#2%

[cl_5y5#1@-3.534];

[cl_5y5#2@1.446];

[cl_5y5#3@-1.833];

[cl_5y5#4@-1.941];

C3 on high_pro;

%c2#3%

[cl_5y5#1@-1.846];

[cl_5y5#2@-0.702];

[cl_5y5#3@2.062];

[cl_5y5#4@-0.993];

C3 on high_pro;

%c2#4%

[cl_5y5#1@-1.658];

[cl_5y5#2@-0.459];

[cl_5y5#3@-0.636];

[cl_5y5#4@2.256];

C3 on high_pro;

%c2#5%

[cl_5y5#1@-12.651];

[cl_5y5#2@-3.724];

[cl_5y5#3@-4.641];

[cl_5y5#4@-5.127];

C3 on high_pro;

MODEL C3:

%C3#1%

[cl_7y#1@13.059];

[cl_7y#2@10.139];

[cl_7y#3 @10.118];

C4 on high_pro;

%C3#2%

[cl_7y#1@-2.691];

[cl_7y#2@1.766];

[cl_7y#3 @-1.356];

C4 on high_pro;

%C3#3%

[cl_7y# 1 @-1.510];

[cl_7y#2@-0.180];

[cl_7y#3@2.249];

C4 on high_pro;

%C3#4%

[cl_7y#1@-13.777];

[cl_7y#2@-3.495];

[cl_7y#3@-4.678];

C4 on high_pro;

MODEL C4:

%C4#1%

[cl_11y#1@12.443];

[cl_11y#2@9.568];

[cl_11y#3@9.422];

C5 on high_pro;

%C4#2%

[cl_11y#1@-2.702];

[cl_11y#2@1.786];

[cl_11y#3 @-1.287];

C5 on high_pro;

%C4#3%

[cl_11y#1@-2.076];

[cl_11y#2@-0.497];

[cl_11y#3@2.156];

C5 on high_pro;

%C4#4%

[cl_11y#1@-12.520];

[cl_11y#2@-4.009];

[cl_11y#3@-4.632];

C5 on high_pro;

MODEL C5: !Notice that in this last part we do not include the covariate.

%c5#1%

[cl_14y5#1@9.107];

[cl_14y5#2@6.174];

[cl_14y5#3@-4.411];

[cl_14y5#4@5.994];

%c5#2%

[cl_14y5#1@-2.940];

[cl_14y5#2@1.869];

[cl_14y5#3@-1.375];

[cl_14y5#4@-2.125];

%c5#3%

[cl_14y5#1@-10.333];

[cl_14y5#2@-1.096];

[cl_14y5#3@1.531];

[cl_14y5#4@-1.757];

%c5#4%

[cl_14y5#1@5.145];

[cl_14y5#2@6.057];

[cl_14y5#3@6.341];

[cl_14y5#4@8.834];

%c5#5%

[cl_14y5#1@-13.774];

[cl_14y5#2@-3.687];

[cl_14y5#3@-4.036];

[cl_14y5#4@-12.505];

Below is an excerpt of the output:

Latent Class Pattern C3#4 ! this means the transition from latent class/profile C3#4 (age 7 Low !symptoms) to age 11 latent profiles, compared to age 11 reference profile (age 11 Low symptoms).

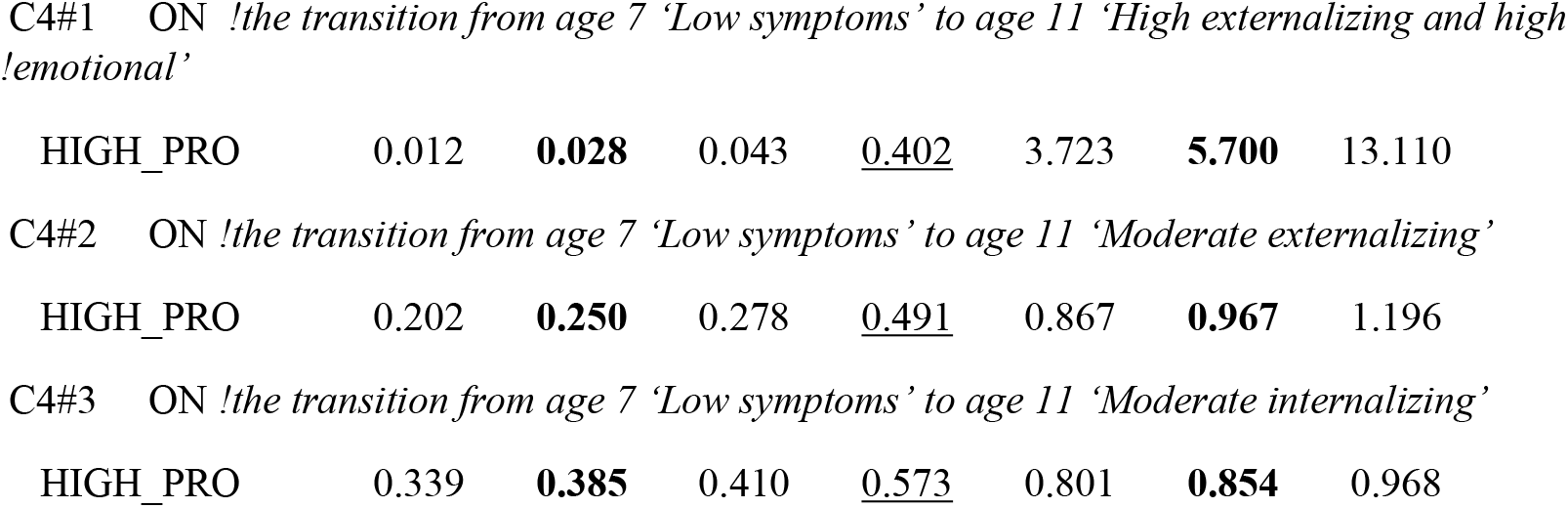

|  |  |  |  |  |  |  |  |
| --- | --- | --- | --- | --- | --- | --- | --- |
| HIGH_PRO | 0.012 | **0.028** | 0.043 | <u>0.402</u> | 3.723 | **5.700** | 13.110 |
| C4#2 ON !the transition from age 7 'Low symptoms' to age 11 'Moderate externalizing' |  |  |  |  |  |  |  |
| HIGH_PRO | 0.202 | **0.250** | 0.278 | <u>0.491</u> | 0.867 | **0.967** | 1.196 |
| C4#3 ON !the transition from age 7 'Low symptoms' to age 11 'Moderate internalizing' |  |  |  |  |  |  |  |
| HIGH_PRO | 0.339 | **0.385** | 0.410 | <u>0.573</u> | 0.801 | **0.854** | 0.968 |

Notice that the middle value is the estimate (underscored), and the bold values are the 95% confidence intervals. Immediately adjacent to the estimates are the 90% confidence intervals. The extreme values are the 99% confidence intervals. (In the original output these values are not highlighted.)

##### F. Association between emotional and behavioural profiles and adolescence mental wellbeing, self-harm, and substance use. (Table 2 in the manuscript)

TITLE: functional outcomes alcohol lifetime

DATA:

FILE IS "*the directory of the datalist*";

TYPE = IMPUTATION;

DEFINE: *! in this part we define the latent profile membership dummy variables*

p1= 0; !p*1 is age 3 High externalizing and moderate emotional*

IF (CL_3Y5 EQ 1) THEN p1 = 1;

IF (CL_3Y5 EQ _MISSING) THEN p1 = _MISSING;

p2= 0; !p*2 is age 3 Moderate externalizing*

IF (CL_3Y5 EQ 2) THEN p2 = 1;

IF (CL_3Y5 EQ _MISSING) THEN p2 = _MISSING;

p3= 0; !*p3 is age 3 Moderate peer problems*

IF (CL_3Y5 EQ 3) THEN p3 = 1;

IF (CL_3Y5 EQ _MISSING) THEN p3 = _MISSING;

p4= 0; *!p4 is age 3 High emotional and conduct*

IF (CL_3Y5 EQ 4000) THEN p4 = 1;

IF (CL_3Y5 EQ _MISSING) THEN p4 = _MISSING;

VARIABLE:

NAMES = ID PTTYPE2 SPTN00 WEIGHT2 emo_3 cond_3 hype_3 peers_3 prosoc_3 emo_5 cond_5 hype_5 peers_5 prosoc_5 emo_7 cond_7 hype_7 peers_7 prosoc_7 emo_11 cond_11 hype_11 peer_11 prosoc_11 emo_14 cond_14 hyper_14 peer_14 prosoc_14 male white harsh_3 harsh_5 harsh_7 pov_9m pov_3 pov_5 pov_7 pov_11 pov_14 high_pro kess_3 kess_5 kess_7 kess_11 kess_14 mono_9m mono_3 mono_5 mono_7 mono_11 mono_14 alc_14 smok_14 binge_14 cann_14 sharm mentw_14 cl_3y5 cl_5y5 cl_3y5 cl_7y cl_11y cl_14y5;

MISSING=*;

IDVARIABLE IS ID;

WEIGHT IS weight2;

STRATIFICATION IS pttype2;

CLUSTER IS sptn00;

categorical= ALC_14; !the outcome variable, in this case alcohol lifetime use at age 14 usevar= male white HARSH_3 POV_9M high_pro KESS_3 MONO_9M ALC_14 p1 p2 p3 p4;

ANALYSIS:

TYPE IS COMPLEX;

Estimator = MLR;

PROCESSORS = 8;

MODEL:

ALC_14 ON male HARSH_3 POV_9M KESS_3 MONO_9M p1 p2 p3 p4 white uni;

OUTPUT:TECH1 sampstat standardized cinterval;

An excerpt of the confidence interval output obtained from this analysis:

CONFIDENCE INTERVALS FOR THE LOGISTIC REGRESSION ODDS RATIO RESULTS

ALC_14 ON

|  |  |  |  |  |  |  |  |
| --- | --- | --- | --- | --- | --- | --- | --- |
| MALE | 0.985 | 1.014 | 1.029 | 1.111 | 1.200 | 1.217 | 1.253 |
| HARSH_3 | 1.021 | 1.028 | 1.031 | 1.050 | 1.069 | 1.072 | 1.079 |
| POV_9M | 0.805 | 0.841 | 0.859 | 0.964 | 1.080 | 1.104 | 1.153 |
| KESS_3 | 0.979 | 0.983 | 0.985 | 0.997 | 1.008 | 1.010 | 1.015 |
| MONO_9M | 1.063 | 1.108 | 1.132 | 1.267 | 1.417 | 1.448 | 1.510 |
| P1 | 0.456 | **0.495** | 0.516 | <u>0.644</u> | 0.804 | **0.839** | 0.911 |
| P2 | 0.888 | **0.918** | 0.934 | <u>1.022</u> | 1.117 | **1.136** | 1.175 |
| P3 | 0.647 | **0.674** | 0.688 | <u>0.767</u> | 0.854 | **0.872** | 0.908 |
| P4 | 0.491 | **0.531** | 0.553 | <u>0.681</u> | 0.838 | **0.873** | 0.943 |
| WHITE | 2.619 | 2.777 | 2.862 | 3.348 | 3.917 | 4.037 | 4.281 |
| HIGH_PRO | 0.786 | 0.810 | 0.822 | 0.890 | 0.963 | 0.977 | 1.006 |

Notice that the middle value is the estimate (underscored), and the bold values are the 95% confidence intervals. Immediately adjacent to the estimates are the 90% confidence intervals. The extreme values are the 99% confidence intervals. (In the original output these values are not highlighted.)G. Testing for measurement invariance, non-invariance and partial measurement invariance

##### G. Testing for measurement invariance, non-invariance and partial measurement invariance

Below we show the syntax to test for measurement non-invariance, partial measurement invariance, and full measurement invariance. After running these models, we obtained the Bayesian information criterion (BIC) and compared the values obtained, with lower values meaning better fit. In these analyses, we did not use the manual three-step specification, as we were only interested in obtaining the BIC.

MEASUREMENT NON-INVARIANCE:

MODEL C1:

%C1#1%
[emo_3 cond_3 hype_3 peers_3 prosoc_3];
%C1#2%
[emo_3 cond_3 hype_3 peers_3 prosoc_3];
%C1#3%
[emo_3 cond_3 hype_3 peers_3 prosoc_3];
%C1#4%
[emo_3 cond_3 hype_3 peers_3 prosoc_3];
%C1#5%
[emo_3 cond_3 hype_3 peers_3 prosoc_3];
MODEL C2:
%C2#1%
[emo_5 cond_5 hype_5 peers_5 prosoc_5];
%C2#2%
[emo_5 cond_5 hype_5 peers_5 prosoc_5];
%C2#3%
[emo_5 cond_5 hype_5 peers_5 prosoc_5];
%C2#4%
[emo_5 cond_5 hype_5 peers_5 prosoc_5];
%C2#5%
[emo_5 cond_5 hype_5 peers_5 prosoc_5];
MODEL C3:
%C3#1%
[emo_7 cond_7 hype_7 peers_7 prosoc_7];
%C3#2%
[emo_7 cond_7 hype_7 peers_7 prosoc_7];
%C3#3%
[emo_7 cond_7 hype_7 peers_7 prosoc_7];[emo_7 cond_7 hype_7 peers_7 prosoc_7];
MODEL C4:
%C4#1%
[emo_11 cond_11 hype_11 peer_11 prosoc_11];
%C4#2%
[emo_11 cond_11 hype_11 peer_11 prosoc_11];
%C4#3%
[emo_11 cond_11 hype_11 peer_11 prosoc_11];
%C4#4%
[emo_11 cond_11 hype_11 peer_11 prosoc_11];
MODEL C5:
%C5#1%
[emo_14 cond_14 hyper_14 peer_14 prosoc_14];
%C5#2%
[emo_14 cond_14 hyper_14 peer_14 prosoc_14];
%C5#3%
[emo_14 cond_14 hyper_14 peer_14 prosoc_14];
%C5#4%
[emo_14 cond_14 hyper_14 peer_14 prosoc_14];
%C5#5%
[emo_14 cond_14 hyper_14 peer_14 prosoc_14];

PARTIAL MEASUREMENT INVARIANCE age 3 = age 5;

MODEL C1:

%C1#1%
[emo_3 cond_3 hype_3 peers_3 prosoc_3] (t1-t5);
%C1#2%
[emo_3 cond_3 hype_3 peers_3 prosoc_3] (tt1-tt5);
%C1#3%
[emo_3 cond_3 hype_3 peers_3 prosoc_3] (ttt1-ttt5);
%C1#4%
[emo_3 cond_3 hype_3 peers_3 prosoc_3] (tttt1-tttt5);
MODEL C2:
%C2#1%
[emo_5 cond_5 hype_5 peers_5 prosoc_5] (t1-t5);
%C2#2%
[emo_5 cond_5 hype_5 peers_5 prosoc_5] (tt1-tt5);
%C2#3%
[emo_5 cond_5 hype_5 peers_5 prosoc_5] (ttt1-ttt5);
%C2#4%
[emo_5 cond_5 hype_5 peers_5 prosoc_5] (tttt1-tttt5);
MODEL C3:
%C3#1%
[emo_7 cond_7 hype_7 peers_7 prosoc_7];
%C3#2%
[emo_7 cond_7 hype_7 peers_7 prosoc_7];
%C3#3%
[emo_7 cond_7 hype_7 peers_7 prosoc_7];
%C3#4%
[emo_7 cond_7 hype_7 peers_7 prosoc_7];
MODEL C4:
%C4#1%
[emo_11 cond_11 hype_11 peer_11 prosoc_11];
%C4#2%
[emo_11 cond_11 hype_11 peer_11 prosoc_11];
%C4#3%
[emo_11 cond_11 hype_11 peer_11 prosoc_11];
%C4#4%
[emo_11 cond_11 hype_11 peer_11 prosoc_11];
MODEL C5:
%C5#1%
[emo_14 cond_14 hyper_14 peer_14 prosoc_14];
%C5#2%
[emo_14 cond_14 hyper_14 peer_14 prosoc_14];
%C5#3%
[emo_14 cond_14 hyper_14 peer_14 prosoc_14];
%C5#4%
[emo_14 cond_14 hyper_14 peer_14 prosoc_14];

PARTIAL MEASUREMENT INVARIANCE age 7 = age 11;

MODEL C1:

%C1#1%
[emo_3 cond_3 hype_3 peers_3 prosoc_3];
%C1#2%
[emo_3 cond_3 hype_3 peers_3 prosoc_3];
%C1#3%
[emo_3 cond_3 hype_3 peers_3 prosoc_3];
%C1#4%
[emo_3 cond_3 hype_3 peers_3 prosoc_3];
%C1#5%
[emo_3 cond_3 hype_3 peers_3 prosoc_3];
MODEL C2:
%C2#1%
[emo_5 cond_5 hype_5 peers_5 prosoc_5];
%C2#2%
[emo_5 cond_5 hype_5 peers_5 prosoc_5];
%C2#3%
[emo_5 cond_5 hype_5 peers_5 prosoc_5];
%C2#4%
[emo_5 cond_5 hype_5 peers_5 prosoc_5];
%C2#5%
[emo_5 cond_5 hype_5 peers_5 prosoc_5];
MODEL C3:
%C3#1%
[emo_7 cond_7 hype_7 peers_7 prosoc_7] (t1-t5);
%C3#2%
[emo_7 cond_7 hype_7 peers_7 prosoc_7] (tt1-tt5);
%C3#3%
[emo_7 cond_7 hype_7 peers_7 prosoc_7] (ttt1-ttt5);
%C3#4%
[emo_7 cond_7 hype_7 peers_7 prosoc_7] (tttt1-tttt5);
MODEL C4:
%C4#1%
[emo_11 cond_11 hype_11 peer_11 prosoc_11] (t1-t5);
%C4#2%
[emo_11 cond_11 hype_11 peer_11 prosoc_11] (tt1-tt5);
%C4#3%
[emo_11 cond_11 hype_11 peer_11 prosoc_11] (ttt1-ttt5);
%C4#4%
[emo_11 cond_11 hype_11 peer_11 prosoc_11] (tttt1-tttt5);
MODEL C5:
%C5#1%
[emo_14 cond_14 hyper_14 peer_14 prosoc_14];
%C5#2%
[emo_14 cond_14 hyper_14 peer_14 prosoc_14];
%C5#3%
[emo_14 cond_14 hyper_14 peer_14 prosoc_14];
%C5#4%
[emo_14 cond_14 hyper_14 peer_14 prosoc_14];
%C5#5%
[emo_14 cond_14 hyper_14 peer_14 prosoc_14];

PARTIAL MEASUREMENT INVARIANCE age 3 = age 5 AND age 7 = age 11;

MODEL C1:

%C1#1%
[emo_3 cond_3 hype_3 peers_3 prosoc_3] (t1-t5);
%C1#2%
[emo_3 cond_3 hype_3 peers_3 prosoc_3](tt1-tt5);
%C1#3%
[emo_3 cond_3 hype_3 peers_3 prosoc_3](ttt1-ttt5);
%C1#4%
[emo_3 cond_3 hype_3 peers_3 prosoc_3](tttt1-tttt5);
%C1#5%
[emo_3 cond_3 hype_3 peers_3 prosoc_3](ttttt1-ttttt5);
MODEL C2:
%C2#1%
[emo_5 cond_5 hype_5 peers_5 prosoc_5] (t1-t5);
%C2#2%
[emo_5 cond_5 hype_5 peers_5 prosoc_5] (tt1-tt5);
%C2#3%
[emo_5 cond_5 hype_5 peers_5 prosoc_5] (ttt1-ttt5);
%C2#4%
[emo_5 cond_5 hype_5 peers_5 prosoc_5] (tttt1-tttt5);
%C2#5%
[emo_5 cond_5 hype_5 peers_5 prosoc_5](ttttt1-ttttt5);
MODEL C3:
%C3#1%
[emo_7 cond_7 hype_7 peers_7 prosoc_7] (a1-a5);
%C3#2%
[emo_7 cond_7 hype_7 peers_7 prosoc_7] (aa1-aa5);
%C3#3%
[emo_7 cond_7 hype_7 peers_7 prosoc_7] (aaa1-aaa5);
%C3#4%
[emo_7 cond_7 hype_7 peers_7 prosoc_7] (aaaa1-aaaa5);
MODEL C4:
%C4#1%
[emo_11 cond_11 hype_11 peer_11 prosoc_11] (a1-a5);
%C4#2%
[emo_11 cond_11 hype_11 peer_11 prosoc_11] (aa1-aa5);
%C4#3%
[emo_11 cond_11 hype_11 peer_11 prosoc_11] (aaa1-aaa5);
%C4#4%
[emo_11 cond_11 hype_11 peer_11 prosoc_11] (aaaa1-aaaa5);

FULL MEASUREMENT INVARIANCE

For this part of the analysis, we had to define four profiles for each age, to be able to test for measurement invariance, which is not possible when the number of profiles differs across transitions.

MODEL C1:

%C1#1%
[emo_3 cond_3 hype_3 peers_3 prosoc_3] (t1-t5);
%C1#2%
[emo_3 cond_3 hype_3 peers_3 prosoc_3] (tt1-tt5);
%C1#3%
[emo_3 cond_3 hype_3 peers_3 prosoc_3] (ttt1-ttt5);
%C1#4%
[emo_3 cond_3 hype_3 peers_3 prosoc_3] (tttt1-tttt5);
MODEL C2:
%C2#1%
[emo_5 cond_5 hype_5 peers_5 prosoc_5] (t1-t5);
%C2#2%
[emo_5 cond_5 hype_5 peers_5 prosoc_5] (tt1-tt5);
%C2#3%
[emo_5 cond_5 hype_5 peers_5 prosoc_5] (ttt1-ttt5);
%C2#4%
[emo_5 cond_5 hype_5 peers_5 prosoc_5] (tttt1-tttt5);
MODEL C3:
%C3#1%
[emo_7 cond_7 hype_7 peers_7 prosoc_7] (t1-t5);
%C3#2%
[emo_7 cond_7 hype_7 peers_7 prosoc_7] (tt1-tt5);
%C3#3%
[emo_7 cond_7 hype_7 peers_7 prosoc_7] (ttt1-ttt5);
%C3#4%
[emo_7 cond_7 hype_7 peers_7 prosoc_7] (tttt1-tttt5);
MODEL C4:
%C4#1%
[emo_11 cond_11 hype_11 peer_11 prosoc_11] (t1-t5);
%C4#2%
[emo_11 cond_11 hype_11 peer_11 prosoc_11] (tt1-tt5);
%C4#3%
[emo_11 cond_11 hype_11 peer_11 prosoc_11] (ttt1-ttt5);
%C4#4%
[emo_11 cond_11 hype_11 peer_11 prosoc_11] (tttt1-tttt5);
MODEL C5:
%C5#1%
[emo_14 cond_14 hyper_14 peer_14 prosoc_14] (t1-t5);
%C5#2%
[emo_14 cond_14 hyper_14 peer_14 prosoc_14] (tt1-tt5);
%C5#3%
[emo_14 cond_14 hyper_14 peer_14 prosoc_14] (ttt1-ttt5);
%C5#4%
[emo_14 cond_14 hyper_14 peer_14 prosoc_14] (tttt1-tttt5);

##### H. R code for the figures in the manuscript

**Latent profile plots (Figure 1):**

library(MplusAutomation)

library(ggplot2)

#LPA age 3#

lpa3 = readModels("insert here the directory of the mplus.out file”)

plpa3 <- plotMixtures(lpa3, rawdata = FALSE, bw=FALSE, ci=NULL)+ scale_color_manual(values=c("#FF6666", "#CC9900", "#33CC66","#3399FF","#CC33CC"))

labels3 <- c("Emotional","Peer","Conduct","Hyperactivity","Prosocial")

plpa3 <- plpa3 + scale_x_discrete(labels= labels3, limits=c("Emo_3", "Peers_3", "Cond_3", "Hype_3", "Prosoc_3"))+ggtitle("Latent profiles age 3 n=14830") + xlab("") + ylab("Mean SDQ score") +scale_y_continuous(breaks=c(0,2,4,6,8,10),limits=c(0,10))

plpa3 <- plpa3 + theme(legend.position="bottom")+ theme(legend.title=element_blank())

#repeat the above code for each age

**Latent transition probabilities (Figure 2):**

library(MplusAutomation)

results = readModels("insert here the directory of the mplus.out file”)

plotLTA(results, node_stroke=3,max_edge_width=4, x_labels = c('C1' = 'Age 3', 'C2'= 'Age 5', 'C3'='Age 7', 'C4'= 'Age 11', 'C5'= 'Age 14'),

> node_labels = c("C1.5"="LS", "C1.4"="He+C", "C1.3"="MPP", "C1.2"="ME", "C1.1"="HE+LP",
>
> "C2.5"="LS", "C2.4"="Me", "C2.3"="MPP", "C2.2"="ME", "C2.1"= "HE+Me",
>
> "C3.4"="LS", "C3.3"="MPP", "C3.2"= "ME", "C3.1"="HE+Me",
>
> "C4.4"="LS", "C4.3"="MI", "C4.2"="ME", "C4.1"= "HE+He",
>
> "C5.5"="LS", "C5.4"="HI", "C5.3"="MI", "C5.2"="ME", "C5.1"="HE+Me"))

**Latent transition analysis with covariates (Figure 3):**

For these figures, one has first to prepare a table for each latent profile, extracting the results from the Mplus outputs. The columns of these tables should have the variable name (term), the estimate, a column for the lower value of the confidence interval (conf.low), and the higher value (conf.high), and the p-value (p.value). For the dotwhisker package to correctly read the tables, the columns should have the labels shown in brackets.

library(dplyr)

library(ggplot2)

library(gridExtra)

library(dotwhisker)

library(broom)

three_help <- read.table(file="*insert here the directory of the previously prepared tab separated file* .txt",header=TRUE, dec=",")

three_me <- read.table(file="*insert here the directory of the previously prepared tab separated file* .txt",header=TRUE, dec=",")

three_mpp <- read.table(file="*insert here the directory of the previously prepared tab separated file* .txt",header=TRUE, dec=",")

three_hec <- read.table(file="*insert here the directory of the previously prepared tab separated file* .txt",header=TRUE, dec=",")

three_help <- three_help %>% mutate(model = "High externalizing + low prosocial") %>% relabel_predictors(c('gender' = "Male",'ethnicity' = "White",'mono' = "Monoparental family",'poverty' = "Poverty",'uni' = "High maternal prof. qual.", 'maternal' = "Maternal distress",'harsh' = "Harsh parenting")) ###this code is to create a new column in the datadrames with the model label, and to change the labels of the variables

three_me <- three_me %>% mutate(model = "Moderate externalizing") %>% relabel_predictors(c('gender' = "Male",'ethnicity' = "White",'mono' = "Monoparental family",'poverty' = "Poverty",'uni' = "High maternal prof. qual.", 'maternal' = "Maternal distress",'harsh' = "Harsh parenting"))

three_mpp <- three_mpp %>% mutate(model = "Moderate peer problems") %>% relabel_predictors(c('gender' = "Male",'ethnicity' = "White",'mono' = "Monoparental family",'poverty' = "Poverty",'uni' = "High maternal prof. qual.", 'maternal' = "Maternal distress",'harsh' = "Harsh parenting"))

three_hec <- three_hec %>% mutate(model = "High emotional + conduct") %>% relabel_predictors(c('gender' = "Male",'ethnicity' = "White",'mono' = "Monoparental family",'poverty' = "Poverty",'uni' = "High maternal prof. qual.", 'maternal' = "Maternal distress",'harsh' = "Harsh parenting"))

model_age_three <- rbind(three_help, three_me, three_mpp,three_hec)

m_three <- dwplot(model_age_three, dodge_size = 0.4, vline = geom_vline(xintercept = 1, colour = "grey60", linetype = 2), dot_args = list(size=2, aes(shape = model)),

whisker_args = list(size=1,aes(linetype = model))) +

theme_bw() + xlab("") + ylab("") + scale_x_log10(breaks=c(0.1,0.5, 1,2,3,5))+

ggtitle("Age 3") +

theme(legend.position = c(0.993, 0.27),

> legend.justification=c(1, 1),
>
> legend.background = element_rect(colour="grey80"),
>
> legend.title = element_blank(),axis.text.x = element_text(size=10),
>
> axis.text.y = element_text(size=10),legend.text=element_text(size=10))
>
> #repeat the code for each age as in figure 3.

## Notes

### Competing Interest Statement

The authors have declared no competing interest.

## References

1. Angold A, Egger HL (2007) Preschool psychopathology: Lessons for the lifespan. J Child Psychol & Psychiat 48:961–966. doi: 10.1111/j.1469-7610.2007.01832.x

2. Luby JL, Gaffrey MS, Tillman R, et al. (2014) Trajectories of preschool disorders to full DSM depression at school age and early adolescence: continuity of preschool depression. Am J Psychiatry 171:768–776. doi: 10.1176/appi.ajp.2014.13091198

3. Roza SJ, Hofstra MB, van der Ende J, Verhulst FC (2003) Stable prediction of mood and anxiety disorders based on behavioral and emotional problems in childhood: a 14-year follow-up during childhood, adolescence, and young adulthood. Am J Psychiatry 160:2116–2121. doi: 10.1176/appi.ajp.160.12.2116

4. Pardini DA, Fite PJ (2010) Symptoms of conduct disorder, oppositional defiant disorder, attention-deficit/hyperactivity disorder, and callous-unemotional traits as unique predictors of psychosocial maladjustment in boys: advancing an evidence base for DSM-V. J Am Acad Child Adolesc Psychiatry 49:1134–1144. doi: 10.1016/j.jaac.2010.07.010

5. Patalay P, Moulton V, Goodman A, Ploubidis GB (2017) Cross-Domain Symptom Development Typologies and Their Antecedents: Results From the UK Millennium Cohort Study. J Am Acad Child Adolesc Psychiatry 56:765–776.e2. doi: 10.1016/j.jaac.2017.06.009

6. Hill AL, Degnan KA, Calkins SD, Keane SP (2006) Profiles of externalizing behavior problems for boys and girls across preschool: The roles of emotion regulation and inattention. Dev Psychol 42:913–928. doi: 10.1037/0012-1649.42.5.913

7. Wichstrøm L, Belsky J, Steinsbekk S (2017) Homotypic and heterotypic continuity of symptoms of psychiatric disorders from age 4 to 10 years: a dynamic panel model. J Child Psychol Psychiatry 58:1239–1247. doi: 10.1111/jcpp.12754

8. Basten M, Tiemeier H, Althoff RR, et al. (2016) The stability of problem behavior across the preschool years: an empirical approach in the general population. J Abnorm Child Psychol 44:393–404. doi: 10.1007/s10802-015-9993-y

9. Bufferd SJ, Dougherty LR, Carlson GA, et al. (2012) Psychiatric disorders in preschoolers: continuity from ages 3 to 6. Am J Psychiatry 169:1157–1164. doi: 10.1176/appi.ajp.2012.12020268

10. University Of London IOE (2019) Millennium Cohort Study: Sixth Survey, 2015. UK Data Service. doi: 10.5255/ukda-sn-8156-4

11. Joshi H, Fitzsimons E (2016) The Millennium Cohort Study: the making of a multi-purpose resource for social science and policy. Longit Life Course Stud. doi: 10.14301/llcs.v7i4.410

12. Goodman A, Goodman R (2009) Strengths and difficulties questionnaire as a dimensional measure of child mental health. J Am Acad Child Adolesc Psychiatry 48:400–403. doi: 10.1097/CHI.0b013e3181985068

13. Collins LM, Lanza ST (2010) Latent class and latent transition analysis: With applications in the social, behavioral, and health sciences. John Wiley & Sons

14. Nylund KL, Asparouhov T, Muthén BO (2007) Deciding on the Number of Classes in Latent Class Analysis and Growth Mixture Modeling: A Monte Carlo Simulation Study. Structural Equation Modeling: A Multidisciplinary Journal 14:535–569. doi: 10.1080/10705510701575396

15. Nylund KL (2007) Latent transition analysis: Modeling extensions and an application to peer victimization. Undergraduate thesis, University of California, Los Angeles Doctoral dissertation

16. Nylund-Gibson K, Grimm R, Quirk M, Furlong M (2014) A Latent Transition Mixture Model Using the Three-Step Specification. Structural Equation Modeling: A Multidisciplinary Journal 21:439–454. doi: 10.1080/10705511.2014.915375

17. Muthén LK, Muthen BO (2017) Mplus Version 8 User’s Guide. 944.

18. Hallquist MN, Wiley JF (2018) MplusAutomation: An R Package for Facilitating Large-Scale Latent Variable Analyses in Mplus. Struct Equ Modeling 25:621–638. doi: 10.1080/10705511.2017.1402334

19. Solt F, Hu Y (2015) dotwhisker: Dot-and-Whisker Plots of Regression Results. The Comprehensive R Archive Network (CRAN)

20. Willner CJ, Gatzke-Kopp LM, Bray BC (2016) The dynamics of internalizing and externalizing comorbidity across the early school years. Dev Psychopathol 28:1033–1052. doi: 10.1017/S0954579416000687

21. Montroy JJ, Bowles RP, Skibbe LE, et al. (2016) The development of self-regulation across early childhood. Dev Psychol 52:1744–1762. doi: 10.1037/dev0000159

22. Wakschlag LS, Tolan PH, Leventhal BL (2010) Research Review: “Ain”t misbehavin’: Towards a developmentally-specified nosology for preschool disruptive behavior. J Child Psychol Psychiatry 51:3–22. doi: 10.1111/j.1469-7610.2009.02184.x

23. Conway AM, McDonough SC (2006) Emotional resilience in early childhood: developmental antecedents and relations to behavior problems. Ann N Y Acad Sci 1094:272–277. doi: 10.1196/annals.1376.033

24. Khan A, McCormack HC, Bolger EA, et al. (2015) Childhood Maltreatment, Depression, and Suicidal Ideation: Critical Importance of Parental and Peer Emotional Abuse during Developmental Sensitive Periods in Males and Females. Front Psychiatry 6:42. doi: 10.3389/fpsyt.2015.00042

25. Adkins DE, Wang V, Elder GH (2009) Structure and Stress: Trajectories of Depressive Symptoms across Adolescence and Young Adulthood. Soc Forces 88:31. doi: 10.1353/sof.0.0238

26. Shevlin M, McElroy E, Murphy J (2017) Homotypic and heterotypic psychopathological continuity: a child cohort study. Soc Psychiatry Psychiatr Epidemiol 52:1135–1145. doi: 10.1007/s00127-017-1396-7

27. Reiss F (2013) Socioeconomic inequalities and mental health problems in children and adolescents: a systematic review. Soc Sci Med 90:24–31. doi: 10.1016/j.socscimed.2013.04.026

28. Fergusson DM, Boden JM, Horwood LJ (2007) Exposure to single parenthood in childhood and later mental health, educational, economic, and criminal behavior outcomes. Arch Gen Psychiatry 64:1089–1095. doi: 10.1001/archpsyc.64.9.1089

29. Sieh DS, Visser-Meily JMA, Meijer AM (2013) The relationship between parental depressive symptoms, family type, and adolescent functioning. PLoS One 8:e80699. doi: 10.1371/journal.pone.0080699

30. Hoffmann JP (2017) Family structure and adolescent substance use: an international perspective. Subst Use Misuse 52:1667–1683. doi: 10.1080/10826084.2017.1305413

31. O’Connor TG, Monk C, Burke AS (2016) Maternal affective illness in the perinatal period and child development: findings on developmental timing, mechanisms, and intervention. Curr Psychiatry Rep 18:24. doi: 10.1007/s11920-016-0660-y

32. Wiggins JL, Mitchell C, Hyde LW, Monk CS (2015) Identifying early pathways of risk and resilience: The codevelopment of internalizing and externalizing symptoms and the role of harsh parenting. Dev Psychopathol 27:1295–1312. doi: 10.1017/S0954579414001412

33. Rajyaguru P, Moran P, Cordero M, Pearson R (2019) Disciplinary parenting practice and child mental health: evidence from the UK millennium cohort study. J Am Acad Child Adolesc Psychiatry 58:108–116.e2. doi: 10.1016/j.jaac.2018.06.033

34. McLaughlin KA, Breslau J, Green JG, et al. (2011) Childhood socio-economic status and the onset, persistence, and severity of DSM-IV mental disorders in a US national sample. Soc Sci Med 73:1088–1096. doi: 10.1016/j.socscimed.2011.06.011

35. Meyrose A-K, Klasen F, Otto C, et al. (2018) Benefits of maternal education for mental health trajectories across childhood and adolescence. Soc Sci Med 202:170–178. doi: 10.1016/j.socscimed.2018.02.026

36. Wills TA, Simons JS, Sussman S, Knight R (2016) Emotional self-control and dysregulation: A dual-process analysis of pathways to externalizing/internalizing symptomatology and positive well-being in younger adolescents. Drug Alcohol Depend 163 Suppl 1:S37–45. doi: 10.1016/j.drugalcdep.2015.08.039

37. Curhan AL, Rabinowitz JA, Pas ET, Bradshaw CP (2019) Informant Discrepancies in Internalizing and Externalizing Symptoms in an At-Risk Sample: The Role of Parenting and School Engagement. J Youth Adolesc. doi: 10.1007/s10964-019-01107-x

## Supplementary materials references

1. Zumbo BD, Gadermann AM, Zeisser C (2007) Ordinal versions of coefficients alpha and theta for likert rating scales. J Mod App Stat Meth 6:21–29. doi: 10.22237/jmasm/1177992180

2. Sulik MJ, Blair C, Greenberg M (2017) Child Conduct Problems across Home and School Contexts: A Person-Centered Approach. J Psychopathol Behav Assess 39:46–57. doi: 10.1007/s10862-016-9564-8

3. Goodman A, Goodman R (2009) Strengths and difficulties questionnaire as a dimensional measure of child mental health. J Am Acad Child Adolesc Psychiatry 48:400–403. doi: 10.1097/CHI.0b013e3181985068

4. Youthinmind Ltd. (2015) Scoring the Strengths & Difficulties Questionnaire for age 4–17. https://www.sdqinfo.com/py/sdqinfo/b3.py?language=Englishqz(UK). Accessed 30 Oct 2019

5. Youthinmind Ltd. (2016) Scoring the Strengths & Difficulties Questionnaire for age 2–4. https://www.sdqinfo.com/py/sdqinfo/b3.py?language=Englishqz(UK). Accessed 30 Oct 2019

6. Patalay P, Moulton V, Goodman A, Ploubidis GB (2017) Cross-Domain Symptom Development Typologies and Their Antecedents: Results From the UK Millennium Cohort Study. J Am Acad Child Adolesc Psychiatry 56:765–776.e2. doi: 10.1016/j.jaac.2017.06.009

7. Patalay P, Fitzsimons E (2016) Correlates of mental illness and wellbeing in children: are they the same? results from the UK millennium cohort study. J Am Acad Child Adolesc Psychiatry 55:771–783. doi: 10.1016/j.jaac.2016.05.019

8. Mazza JRSE, Lambert J, Zunzunegui MV, et al. (2017) Early adolescence behavior problems and timing of poverty during childhood: A comparison of lifecourse models. Soc Sci Med 177:35–42. doi: 10.1016/j.socscimed.2017.01.039

9. Hernández-Alava M, Popli G (2017) Children’s development and parental input: evidence from the UK millennium cohort study. Demography 54:485–511. doi: 10.1007/s13524-017-0554-6

10. Kessler RC, Barker PR, Colpe LJ, et al. (2003) Screening for serious mental illness in the general population. Arch Gen Psychiatry 60:184–189. doi: 10.1001/archpsyc.60.2.184

11. Straus MA, Hamby SL (1997) Measuring Physical & Psychological Maltreatment of Children with the Conflict Tactics Scales.

12. Flouri E, Midouhas E (2017) Environmental adversity and children’s early trajectories of problem behavior: The role of harsh parental discipline. J Fam Psychol 31:234–243. doi: 10.1037/fam0000258

13. Plewis I (2007) Non-response in a birth cohort study: the case of the millennium cohort study. Int J Soc Res Methodol 10:325–334. doi: 10.1080/13645570701676955

## Data References

1. University Of London IOE. Millennium Cohort Study: Longitudinal Family File, 2001–2015. UK Data Service. 2017. doi:10.5255/ukda-sn-8172-2

2. University Of London IOE. Millennium Cohort Study: First Survey, 2001–2003. UK Data Service. 2017. doi:10.5255/ukda-sn-4683-4

3. University Of London IOE. Millennium Cohort Study: Second Survey, 2003–2005. UK Data Service. 2017. doi:10.5255/ukda-sn-5350-4

4. University Of London IOE. Millennium Cohort Study: Third Survey, 2006. UK Data Service. 2017. doi:10.5255/ukda-sn-5795-4

5. University Of London IOE. Millennium Cohort Study: Fourth Survey, 2008. UK Data Service. 2017. doi:10.5255/ukda-sn-6411-7

6. University Of London IOE. Millennium Cohort Study: Fifth Survey, 2012. UK Data Service. 2017. doi:10.5255/ukda-sn-7464-4

7. University Of London IOE. Millennium Cohort Study: Sixth Survey, 2015. UK Data Service. 2019. doi:10.5255/ukda-sn-8156-4

